# EHR-ML: A generalisable pipeline for reproducible clinical outcomes using electronic health records

**DOI:** 10.1101/2024.03.02.24302664

**Authors:** Yashpal Ramakrishnaiah, Nenad Macesic, Geoffrey I. Webb, Anton Y. Peleg, Sonika Tyagi

## Abstract

The healthcare landscape is experiencing a transformation with the integration of Artificial Intelligence (AI) into traditional analytic workflows. However, this advancement encounters challenges due to variations in clinical practices, resulting in a crisis of generalisability. Addressing this issue, our proposed solution, EHR-ML, offers an open-source pipeline designed to empower researchers and clinicians. By leveraging institutional Electronic Health Record (EHR) data, EHR-ML facilitates predictive modelling, enabling the generation of clinical insights. EHR-ML stands out for its comprehensive analysis suite, guiding researchers through optimal study design, and its built-in flexibility allowing for construction of robust, customisable models. Notably, EHR-ML integrates a dedicated two-layered ensemble model utilising feature representation learning. Additionally, it includes a feature engineering mechanism to handle intricate temporal signals from physiological measurements. By seamlessly integrating with our quality assurance pipelines, this utility leverages its data standardization and anomaly handling capabilities.

Benchmarking analyses demonstrate EHR-ML’s efficacy, particularly in predicting outcomes like inpatient mortality and the Intensive Care Unit (ICU) Length of Stay (LOS). Models built with EHR-ML outperformed conventional methods, showcasing its generalisability and versatility even in challenging scenarios such as high class-imbalance.

We believe EHR-ML is a critical step towards democratising predictive modelling in health-care, enabling rapid hypothesis testing and facilitating the generation of biomedical knowledge. Widespread adoption of tools like EHR-ML will unlock the true potential of AI in healthcare, ultimately leading to improved patient care.

## 1. Introduction

Artificial Intelligence (AI) is rapidly transforming healthcare. AI-enabled predictive modelling of patient outcomes [1, 2, 3, 4], can support early disease detection, more targeted therapies, and improved risk stratification. Electronic Health Records (EHRs) paves the way for powerful predictive modelling [5, 6, 7, 8, 9]. However, concern around non-generalizability of research outcomes is a recurring theme in EHR-based predictive modelling [10, 11]. Models trained in one centre may not be applicable in other settings due to difference in clinical practices and data characteristics [12, 13]. Hence, to unlock the true potential of AI in healthcare, site-specific modelling is essential, leveraging localised data within the institutional EHRs.

While numerous studies leverage EHRs in this way, they are impaired by the absence of well-established pre-processing techniques [14, 15, 16], modelling tools [17, 18, 19], and protocols [15, 18, 20]. This fragmented landscape leads to irreproducible results, inconsistent outcomes, needless complexity, and error-prone ad-hoc processes [17, 21, 22]. To establish standardization in this process, various frameworks[23, 24] and guidelines [25, 26, 27, 28, 29] are proposed for conducting and reporting such studies. Additionally, generic toolkits for predictive modeling have been developed to accelerate AI adoption in healthcare[30, 31, 32, 6]. However, these toolkits often suffer from some significant limitations. Restricting the data source to a single rigid input format [31, 32], hinders their applicability, while non-interpretable neural networks raise implementation concerns in healthcare settings [30, 6]. Furthermore, existing solutions lack end-to-end automation, covering data sourcing to model building, posing a roadblock for widespread localized deployments. Moreover, limitations such as restrictions in selecting a target, manual feature selection processes, and non-robust performance metrics compromise their utility. Additionally, generic out-of-the-box models offered by these frameworks often underperform compared to models customized with domain-specific nuances. Furthermore, all these approaches require extensive configuration regarding study design choices, including data window selection, target viability assessment, sample size optimization, and pre-processing steps.

Developing an effective study design to construct robust predictive models from EHR data is challenging. One major hurdle involves determining the appropriate time window for data collection, ranging from early-stage predictions within 12-48 hours [33, 34, 35] to encompassing the entire duration of admission [36],or retrospective analysis [37, 38]. Additionally, the scarcity of examples within certain data classes can impede the modeling of specific clinical targets [39, 40]. For example, while predicting outcomes for hospital stays exceeding seven days may be feasible, it becomes impractical for stays surpassing 30 days due to insufficient data samples. Moreover, healthcare data is often constrained by privacy concerns and the high cost [41, 42]. Understanding the minimum data requirement for reliable modeling is crucial. Another challenge stems from the class imbalance, resulting in skewed outcome distributions, known as representation bias. This imbalance, where the class of interest may have fewer instances (e.g., longer hospital stays or patient deaths), can impede model training and necessitate careful data balancing strategies [43, 44]. Additionally, the varied scales of clinical attributes pose obstacles for machine learning models. Attributes like temperature, measured in degrees Celsius, span a range of 35 to 40, while heart rate, measured in beats per minute, typically falls between 60 and 100. Each attribute operates on distinct units and scales. To mitigate this issue, data harmonization and scaling techniques are employed to standardize all variables, benefiting certain modeling approaches. However, the decision to standardize data adds complexity to the modeling process [45]. Furthermore, EHR measurements are recorded at irregular intervals, making it challenging to design data transformation methods that retain both magnitude and temporal dynamics for machine learning models. Making informed choices about these parameters is crucial for successful modeling but currently relies heavily on empirical guesswork due to a lack of appropriate tool sets for exploring optimal parameter values [14, 46].

In response to these challenges, we introduce EHR-ML, a comprehensive package for the predictive modelling of clinical outcomes using the EHR. This package ensures that every stage, from data acquisition to model construction, adheres to a clearly defined, domain-specific, data-centric, and reproducible protocol, enforcing optimal practices. To demonstrate the effectiveness of EHR-ML, we employed it to forecast clinical outcomes within a selected cohort of patients diagnosed with sepsis, a condition demanding time-sensitive intervention and treatment. Within this cohort, we conducted predictive modeling for two key clinical outcomes: patient mortality and Intensive Care Unit (ICU) Length of Stay (LOS) [47, 48, 49].

To demonstrate its utility, we performed predictive analysis for mortality risk, a pivotal area of focus [50, 51, 52, 53], owing to its significance in patient care and resource management. Prompt identification of mortality risk equips healthcare professionals with crucial insights for patient triage, treatment strategies, efficient resource distribution, and a comprehensive comprehension of the factors affecting patient outcomes. Traditionally, severity scoring systems like SOFA [54], qSOFA [55], SAPS II [56], and APACHE [57] have played a crucial role in this regard. However, leveraging localised data, machine learning has emerged as a promising alternative outperforming the one-size-fits-all conventional scoring schemes. The next prediction task deals with forecasting another clinical outcome, the ICU-LOS. The LOS prediction is usually approached as a binary prediction problem [58, 59, 60], although some studies adopt continuous regression modelling methods [61, 62]. Notably, the prediction of LOS poses increased complexity compared to mortality prediction [63], as patient distinctions are less pronounced between the classes in this case. By modelling these two diverse outcomes, we aim to showcase the utility, versatility, and simplicity of EHR-ML.

In essence, the goal of EHR-ML is to bridge existing gaps by providing a user-friendly, open-source platform that enables clinicians and researchers to effectively leverage the potential of institutional healthcare data.

## 2. Methods

### 2.1. Data

The development and assessment of EHR-ML utilised two openly accessible, EHR datasets: Medical Information Mart for Intensive Care (MIMIC) IV [64] and eICU [65]. MIMIC IV consists of the data from a single large tertiary teaching hospital, while eICU includes data from a network of critical care units. From these two sources, three distinct cohorts focused on sepsis patients were extracted (refer to Table M1). From each cohort, vital signs and laboratory measurements were extracted for analysis. Next, both datasets were standardised to the OMOP-CDM schema [66, 67] and mapped to standard SNOMED vocabulary [68]. This was achieved through the standardisation module within our previously published EHR-QC tool [69]. Standardisation facilitates consistent data interpretability, leveraging existing tools, interoperability of developed tools, standardised pre-processing, and deduplication of the data.

### 2.2. Machine learning

For machine learning, the chosen data cohort underwent rigorous preprocessing with the EHR-QC quality assurance module (see supplementary figure S1). Initially, vital signs and laboratory measurements for the patient cohort were extracted. Subsequently, a subset of measurements recorded for a high proportion of patients (over 80% in this work) was retained after coverage analysis. This analysis report lists attributes by prevalence in the EHR, aiding in determining a suitable threshold. The measurements that are rare in the EHR lack sufficient numbers for effective modelling, hence removed from the analysis. Essentially, this process retains the most widely recorded measurements (e.g., temperature and heart rate) whereas, rarely occurring measurements (e.g., Intracranial pressure) are removed.

The subsequent phase entails formatting the chosen data to render it compatible with machine learning tasks. EHR data usually contains high-resolution vital signs and laboratory measurements with many recordings within a short period. While this information is valuable in modelling, directly utilising such data can be a challenge with the feature-based machine learning algorithms that necessitate the data to be structured as a two-dimensional matrix. This is addressed by employing a multi-faceted aggregation approach [70] using five functions: minimum, maximum, first value, last value, and mean. These aggregations are applied to vitals and lab measurements, producing 10 feature sets. Grouping attributes prevents an excessive number of features, mitigating the curse of dimensionality. Each group captures distinct time-series aspects, allowing extraction of statistical features and temporal dynamics. This ensures retention of crucial information on central tendency and variability post-aggregation, resulting in a richer representation for analysis and modeling.

The formatted data typically includes some percentage of missing values, representing unrecorded measurements on specific dates. EHR-QC’s advanced imputation module automatically selects the most suitable method for each data type and missing proportion, providing optimal estimates for missing values. Following this, an unsupervised outlier detection algorithm [71] is employed to identify and remove highly eccentric data points. Ultimately, the QA process produces a high-quality data matrix suitable for modeling purposes.

The quality-assured data encompass all the available data from the patient timeline, whereas there is a need to restrict the data within a certain time frame or time window for modelling purposes. For instance, while using retrospective data for predicting the patient’s risk of mortality post 2 days of admission time, all the futuristic data at the time of prediction (48 hours) needs to be purged.

A patient enters the hospital, marking the start of the analysed timeline denoted as Day 1. If transferred to ICU, the day of ICU admission is marked as Day A and day of discharge marked as Day D. The time-window usually centres around ICU admission (Day A), but can be anchored to any other relevant time point such as the time of positive microbiology culture result. We determine the starting point of our data window with a parameter called “window before” (WB), indicating how much historical data we capture before the anchor time. Conversely, the parameter “window after” (WA) determines the end point of the data window, representing the duration of data considered after the anchor time. The data window thus spans from A - WB to A + WA, covering a total duration of WB + WA. Selecting appropriate values for WA and WB depends on the study design. Increasing WB includes more historical data for modeling, while increasing WA extends the data collection period post anchor point.

The next step is to calculate the target variable for the prediction. In this work, we specifically looked at two clinical outcomes, discharge mortality and ICU LOS. The mortality on discharge is a binary outcome indicating whether the patient survived during the ICU stay. The LOS is calculated as the difference between discharge and admission times (D - A). In this work, to facilitate binary classification, we framed LOS prediction as the probability of a patient exceeding a set hospital stay duration threshold. Specifically, we set the threshold as 7 and 14 days. This approach allows flexible adaptation of the target variable, enabling predictions for any threshold duration. In fact, EHR-ML provides the flexibility to model any clinical outcome of interest that is either directly present or derived from the EHR.

Internally, EHR-ML leverages a two-layer ensemble architecture for robust clinical outcome prediction (Figure 1-B). At level 1, four distinct models - XGBoost (XGB), Logistic Regression (LR), Light Gradient Boosted Machines (LGBM), and Multilayer Perceptron (MLP) – are individually trained on 10 feature sets. As each feature set is used by all four models, a total of 40 models are built at level 1. The predictions from Level 1 models are used as input to the second layer (Level 2). Using the outputs from level 1 models as inputs for level 2 helps to combine the strengths of individual models. This intermediate data, shown in figure 2 to have better discriminative power, serves as input for the final XGBoost prediction. In this architecture, obtaining the important features of the final model will help in understanding factors affecting the clinical outcome under consideration making it interpretable. Hyperparameters for each model at both layers are individually optimised specifically for the corresponding data input, ensuring optimal performance.

**Figure 1:**
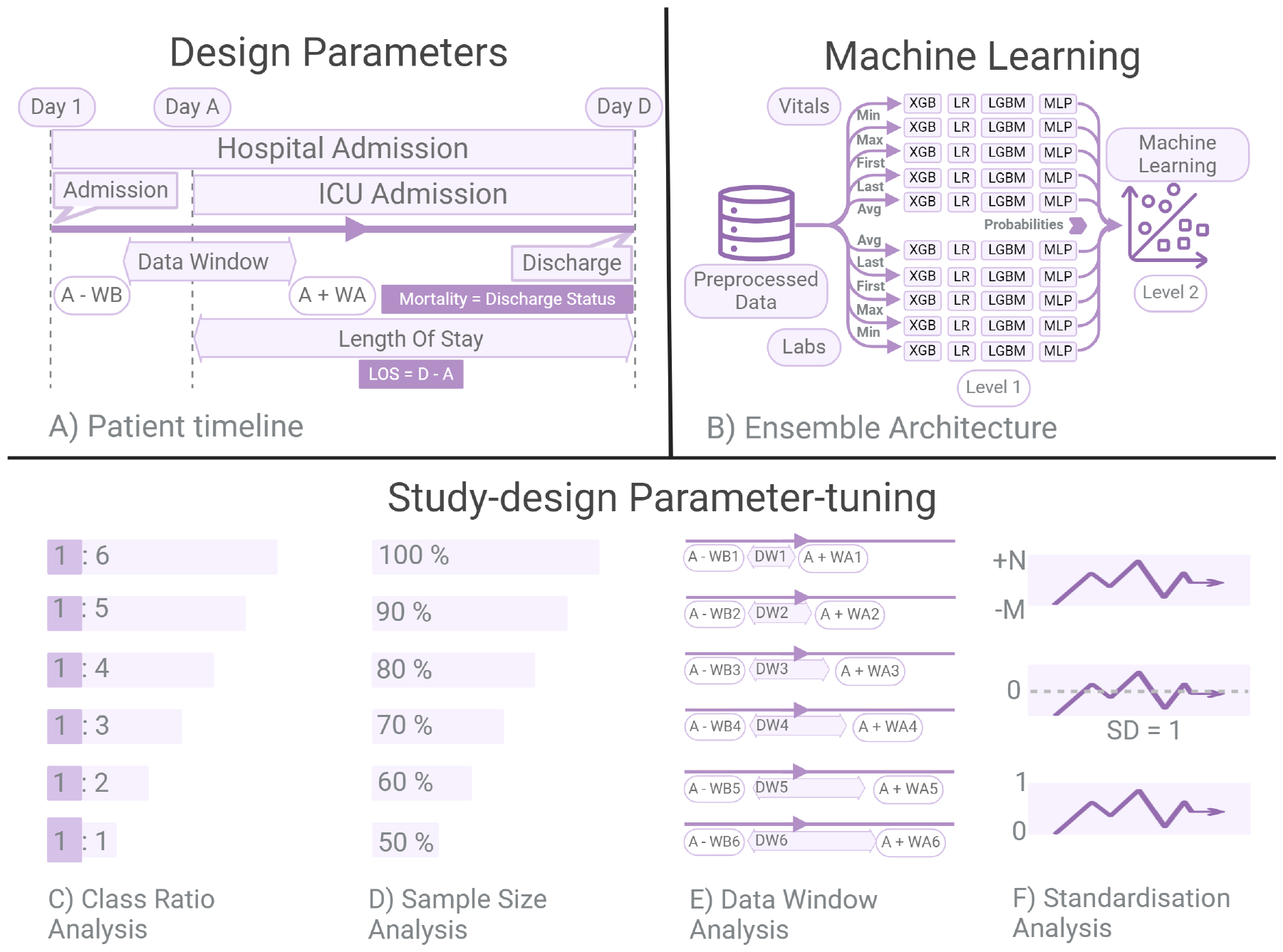
A) This panel depicts a typical patient timeline with key timestamps: hospital admission (Day 1), ICU admission (Day A), and discharge (Day D). The, WB and WA parameters define the data window used for obtaining the data from A - WB to A + WA. The relevant clinical outcomes, LOS [LOS = discharge time D - admission time A], and mortality indicating the patient status at discharge, are also shown. B) This panel illustrates the multi-level ensemble architecture utilised by EHR-ML for machine learning. Level 1 consists of four individual models – XGB, LR, LGBM, and MLP – each trained on vital signs and laboratory measurements formed using five different aggregation functions (see methods). Level 2 then combines the learned feature representations to build the final ensemble model. C) This panel depicts datasets with varying positive and negative class ratios, constructed for analysing the impact of class imbalance on model performance. D) This panel showcases datasets of different sizes obtained through sampling, used for analysing the influence of sample size on model performance. For this analysis, both the classes are randomly sampled without any stratification. E) This panel illustrates multiple patient timelines with different data windows generated by varying the WB and WA parameters, used for analysing the influence of data collection window on model performance. F) This panel compares three data representations: raw data without any transformations, data scaled using Standard Scaling (mean of 0 and standard deviation of 1), and data scaled using MinMax Scaling (minimum of 0 and maximum of 1). These variations enable analysis of the impact of data standardisation on model performance.

**Figure 2:**
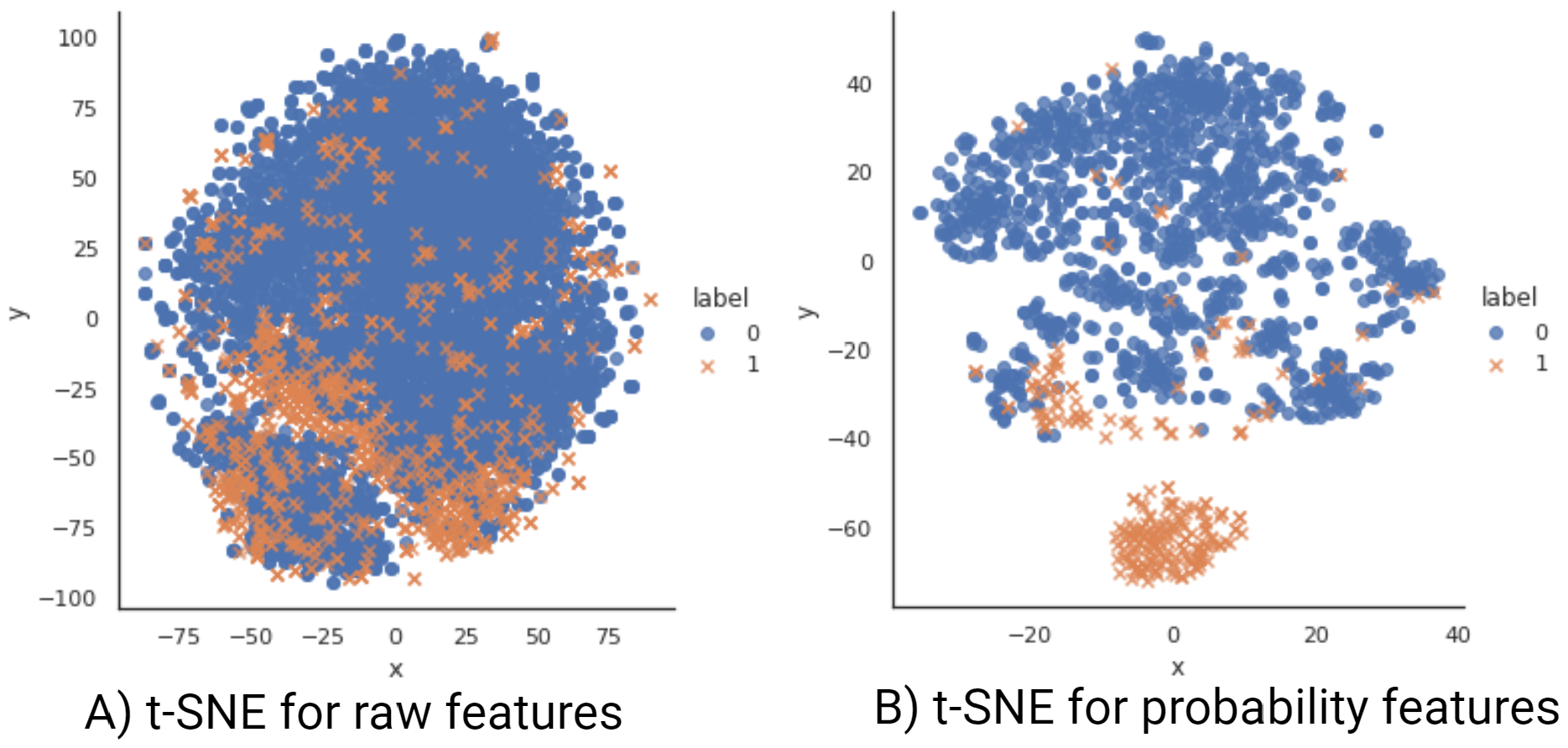
This figure compares the original data (A) to the learned feature representations (B). While the data is high-dimensional and difficult to visualise, t-SNE projects it onto a 2D space for comparison. Each data point is coloured and shaped according to its class (orange and “x” for class 1, blue and “o” for class 0). Comparing the two plots reveals that the learned feature representations lead to a clearer separation between classes. This improved class separability translates to better predictive performance in downstream modelling tasks.

### 2.3. Analytic evaluation and validation

We evaluated and compared the performance of different models using a comprehensive set of metrics and visualisations. For individual model assessment, standard metrics like accuracy, balanced accuracy, average precision, F1 score, area under the receiver operating characteristic curve (AUROC), and Matthews correlation coefficient-F1 (MCCF1) were computed. To facilitate comparisons between different models, we reported True Positives (TP), True Negatives (TN), False Positives (FP), and False Negatives (FN), along with a heatmap of the confusion matrix to visualise them. All metrics were calculated over 5-fold cross-validation, and mean values were reported.

We compared the performance of our ensemble model by benchmarking it against the traditional SAPS II scoring system [56]. SAPS II utilises 12 physiological and 3 disease-related variables, assigning individual scores for variable ranges and aggregating them to calculate a final score. This score can then be used to estimate the risk of mortality through a predefined formula. In addition, we built another model, EHR-ML-SAPS-II, utilising EHR-ML ensemble architecture, but restricting the features to only those used in SAPS-II. This allowed for a direct comparison between the ensemble model’s architecture and traditional scoring methods while controlling for data variations.

### 2.4. Parameter optimisation for study design

EHR-ML encompasses an analysis suite to determine the optimal study design for modelling a specific clinical outcome and guide efficient data collection and preprocessing strategies.

- **Class Ratio Analysis** This analysis explores the fluctuation in model performance as the proportions of positive and negative classes vary (Figure 1-C). Understanding the impact of class imbalance helps identify reliable performance metrics in extreme class ratio scenarios and informs strategies to mitigate the bias. Additionally, it helps assess whether a particular clinical outcome has adequate representation from the minority class to be considered suitable for modeling.
- **Sample Size Analysis** This analysis serves to pinpoint the ideal data size necessary for a specific predictive task, a critical aspect for both retrospective and prospective studies. The process entails randomly sampling data of various sizes and constructing machine learning models utilizing 5-fold cross-validation By doing so, it enables the evaluation of current data sufficiency and provides direction for data augmentation in retrospective studies. Furthermore, it offers insights into sample size necessities for prospective studies. (Figure 1-D).
- **Data Window Analysis** By varying the “window before” (WB) and “window after” (WA) parameters (Figure 1-E), EHR-ML finds the optimal window for collecting data relevant to a prediction task. The best WB parameter determines the sufficient extent of historical data needed, while the best WA parameter reveals the optimal time duration after the admission (or a custom anchor point) for obtaining the data to get reliable outcome predictions.
- **Data Standardisation Analysis** This analysis compares the performance of models trained on raw and scaled data (Figure 1-F). It helps to decide if scaling is beneficial and, if so, which scaling strategy provides the best results. While some machine learning models handle rescaling internally, others are sensitive to it, necessitating careful analysis.

Additionally, EHR-ML offers flexibility by allowing these analyses to be applied either to the ensemble model or a standalone machine learning model.

## 3. Results

### 3.1. EHR-ML outperforms off-the-Shelf models in a comprehensive evaluation

We compared the EHR-ML pipeline against both constituent models within the ensemble and standalone models constructed with standard Python libraries. First, we trained EHR-ML to predict the risk of mortality in patients diagnosed with sepsis [64]. Subsequently, we performed 5-fold cross-validation and obtained the average AUROC, MCCF1, Accuracy, Balanced Accuracy, Average Precision, and F1 scores. These metrics were also calculated for the constituent models within EHR-ML, grouped into four categories based on their machine learning algorithm namely XGB, LR, LGBM, and MLP. Figure 3-A presents a spider plot comparing the performance of EHR-ML with the best-performing constituent model under each category. Further, we trained standalone XGB and LR models on the same dataset. Eventually, the performance of each model was evaluated using various metrics, visualised in figure 3-B and further detailed in supplementary tables S2 and S1.

**Figure 3:**
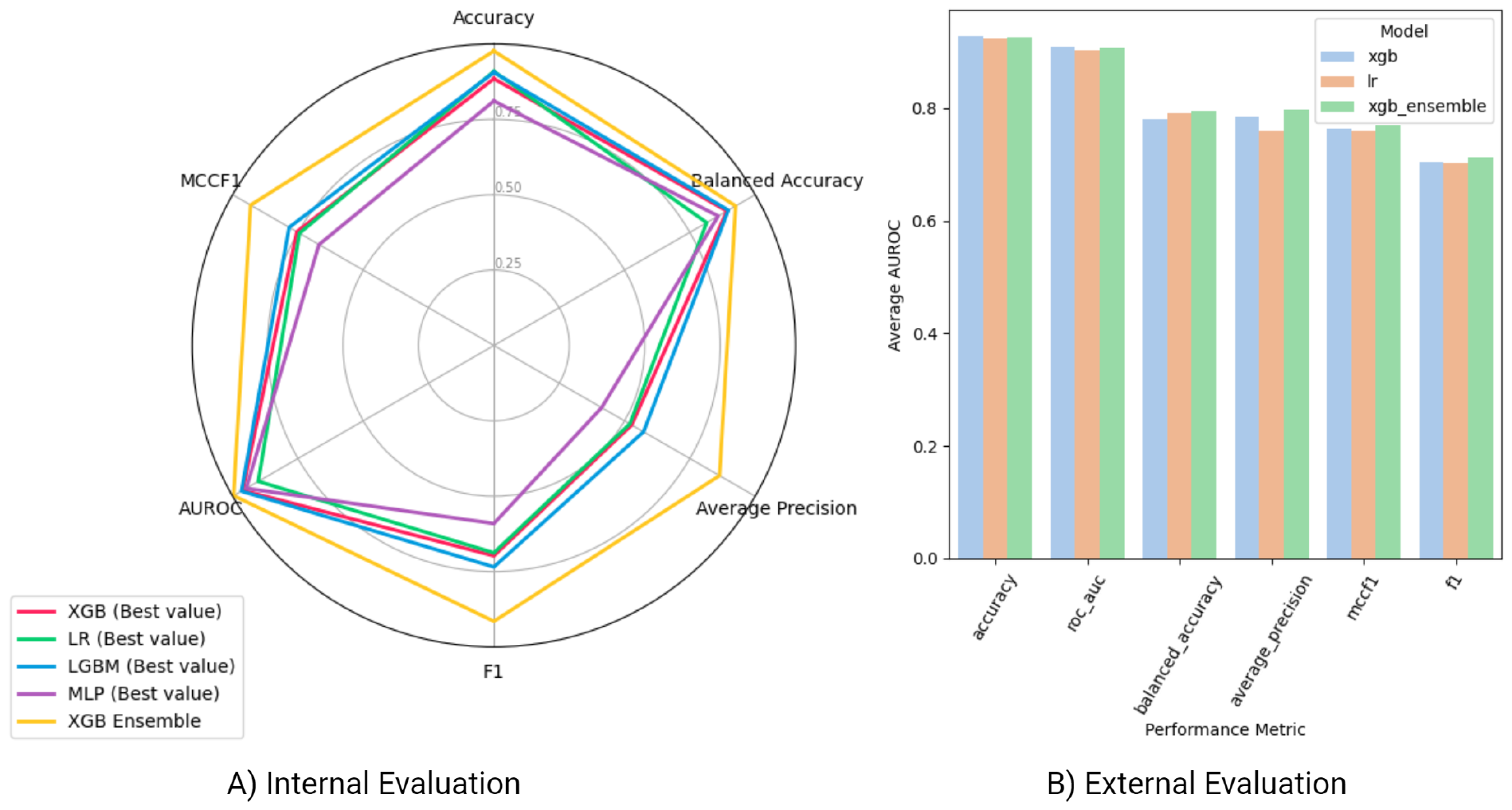
Performance comparison of EHR-ML and individual model constituents across six evaluation metrics, including AUROC, MCCF1, Accuracy, Balanced Accuracy, Average Precision, and F1. A) Panel A compares EHR-ML against the best performing constituent model within each category. The categories include XGB, LR, LGBM, and MLP and there will be 12 individual models under each of them. B) Panel B compares EHR-ML against standalone models trained on the same dataset. Overall, these plots demonstrate the consistent performance advantage of EHR-ML relative to both internal constituent models and external standalone models.

Across both comparative evaluations, EHR-ML consistently demonstrates an improved overall performance against all constituent and standalone models, demonstrating its superior overall performance in clinical prediction tasks. This suggests that the two-layered ensemble approach of EHR-ML successfully leverages the complementary strengths of multiple constituent models to achieve superior performance and generalizability compared to off-the-shelf solutions. Specifically, it benefits as each Level 1 model captures different aspects of the data, leading to a more comprehensive representation upon combining. The well-separated predictions from Level 1 make the final model less susceptible to class imbalance issues. As the second layer operates on probability outputs from the previous layer, the need for data scaling or standardisation is less crucial. Overall, this two-layered architecture allows EHR-ML to extract rich learned feature representations from the data, leverage diverse modelling approaches, and mitigate the impact of class imbalance, ultimately leading to robust and accurate clinical outcome predictions.

### 3.2. EHR-ML outperforms traditional scoring system (SAPS II)

We further evaluated the performance of EHR-ML trained on all the attributes, referred to here as EHR-ML-FULL, against a traditional severity of illness scoring scheme, SAPS II [56]. In addition, we included EHR-ML-SAPS-II, a model also based on EHR-ML architecture but trained only on SAPS II variables. Figure 4-A presents overlapping ROC curves for SAPS-II, EHR-ML-SAPS-II, and EHR-ML-FULL, allowing for comparison of their predictive power across various thresholds. Further, figures 4-B, 4-C, and 4-D further depict the confusion matrices for these models at a calibrated threshold. Furthermore, the table 2 presents their TP, TN, FP, and FN values. This analysis was performed to facilitate the comparison of the strengths and weaknesses of EHR-ML and the SAPS-II in identifying true and false predictions for the two target classes.

**Table 1.** Details of the three patient cohorts used in this study: MIMIC IV ICD (identified by ICD codes), MIMIC IV Micro (microbiologically confirmed sepsis), and eICU ICD (identified by ICD codes). The number of patients and episodes in each cohort, along with the source of the data, inclusion criteria, and the specific condition used for inclusion is presented in this table.

| Cohort | Patients | Episodes | Data Source | Inclusion Criteria | Inclusion Condition |
| --- | --- | --- | --- | --- | --- |
| MIMIC IV ICD | 691 | 729 | MIMIC IV | Sepsis ICD codes | Recorded ICD-9 codes of 995.91, 995.92, and 785.52, and ICD-10 codes of A419, R6520, and R6521 corresponding to sepsis, severe sepsis, and septic shock respectively |
| MIMIC IV Micro | 1790 | 1790 | MIMIC IV | Positive blood culture result | Microbiology investigation yielded a positive result with a bacteria detected |
| eICU ICD | 9612 | 11146 | eICU | Sepsis ICD codes | Recorded ICD-9 codes of 995.91, 995.92, and 785.52 corresponding to sepsis, severe sepsis, and septic shock respectively |

**Table 2.** The table provides a comprehensive summary of performance metrics, including AUROC, TP, TN, FP, and FN for SAPS-II, EHR-ML trained on SAPS II variables - EHR-ML-SAPS-II, and EHR-ML trained on all the variables - EHR-ML-FULL. Higher values are desirable for TP, TN, and AUROC, while lower values are preferred for FP and FN. It can be seen that the EHR-ML trained on full data displays highest performance across all the assessed metrics hence the corresponding row is highlighted.

| Model | AUROC | TP | TN | FP | FN |
| --- | --- | --- | --- | --- | --- |
| SAPS II | 0.5907 | 25.2 % | 92.9 % | 74.8 % | 7.1 % |
| EHR-ML-SAPS-II | 0.7040 | 48.1 % | 92.7 % | 51.7 % | 7.3 % |
| **EHR-ML-FULL** | **0.7217** | **51.4 %** | **93.0 %** | **48.6 %** | **7.0 %** |

**Figure 4:**
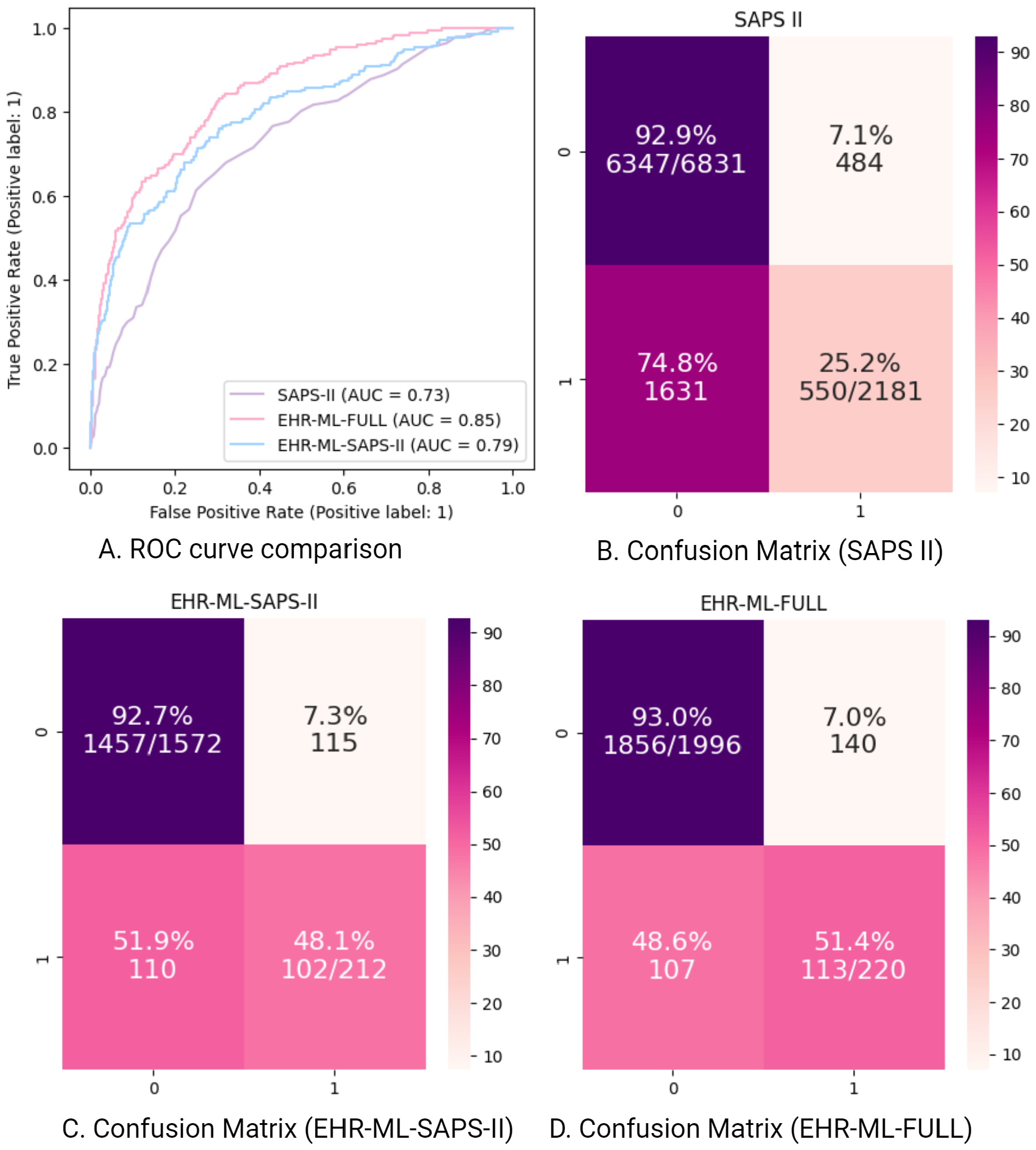
The figure depicting a comparative analysis between the three methods - EHR-ML-FULL, the traditional scoring scheme - SAPS II, and the EHR-ML-SAPS-II model. A) This figure shows an overlapping ROC plot for all three models-EHR-ML-FULL, EHR-ML-SAPS-II, and SAPS-II. B-D) The confusion matrix is presented as a heatmap for SAPS-II, EHR-ML trained on SAPS II variables - EHR-ML-SAPS-II, and EHR-ML trained on all the variables - EHR-ML-FULL. In the confusion matrix, the vertical axis shows actual patient survival while the horizontal axis displays predicted outcomes. Both axes use “0” for the negative class (survival), and “1” for the positive class (deceased). Each cell reveals the number and percentage of patients falling into each category, with the total class counts displayed in the diagonal cells. The variations in colour intensity visually highlights the percentage of observations in each cell.

Figure 4-A reveals the superiority of both EHR-ML variants in predicting mortality risk as compared to conventional SAPS II scoring scheme across various thresholds. Further analysis of the confusion matrices presented in figures 4-B, 4-C, and 4-D reinforces this observation. In all three plots, both EHR-ML variants outperform SAPS II across all quadrants, showing a notable twofold increase in true positives while maintaining similar levels of false positives. This translates to a remarkable improvement, with EHR-ML variants correctly identifying mortality rates twice as often as SAPS II. Moreover, the comparison serves as compelling evidence for the effectiveness of EHR-ML’s two-tier ensemble architecture and the encompassing feature representation learning. The fact that both the models utilise the same data set, highlights the inherent advantage of the EHR-ML’s novel architecture. In conclusion, these findings demonstrate that EHR-ML offers significant improvements over traditional methods like SAPS II in terms of accurately identifying high-risk patients.

### 3.3. Superior performance without the need for scaling

To investigate the requirement of data scaling, we evaluated the model on three different datasets. The raw dataset contained the original, non-scaled features directly extracted from the EHR. Secondly, the standard-scaled data, utilised a standard scaler to transform each feature to have a mean of 0 and a standard deviation of 1. Third, the min-max scaled data, employed a min-max scaler to transform each feature to have a minimum value of 0 and a maximum value of 1. On these three datasets, we performed a 5-fold cross-validation for each dataset to assess the model’s performance in predicting in-hospital mortality (supplementary table S3). Next, we calculated the AUROC for each fold and subsequently generated a box plot summarising the distribution of these values over 5-fold cross-validation (supplementary figure S2).

The EHR-ML model proposed shows excellent accuracy in predicting mortality risk, and it achieves this without needing data scaling. This is because the model’s initial layer generates feature representations that are inherently uniform, with each subset of features producing a risk score between 0 and 1. These scores act as inputs for the second layer of the model, which combines them to make final predictions. Because the inputs to the final model are already within a consistent range, traditional scaling methods are unnecessary. This is a significant advantage of EHR-ML, as it simplifies the model pipeline and reduces computational overhead. Additionally, the absence of data scaling ensures that the model’s predictions are not influenced by scaling parameters, making them more robust and generalizable across different datasets.

### 3.4. EHR-ML excels with both narrow and extended data windows

To determine the amount of data required for the model to make reliable predictions, we performed a data window analysis. The analysis involved varying the size of a temporal window around the anchor point by modifying two parameters namely window before and window after. In this analysis, the window before parameter spanned from 0 to 3 days before the anchor point. Similarly, the window after parameter ranged from 1 to 14 days after the anchor point at the end of which the predictions were made. For each combination of the two parameters, we calculated the EHR-ML model’s performance in terms of average AUROC over 5-fold. These values are tabulated in table 3 and plotted as a heatmap in figure 5, providing a visual representation of the impact of the data window in predicting in-hospital mortality.

**Table 3.** The table presents average AUROC values across various time windows. The data collection window’s lower boundary begins 3 days before the anchor day (ICU admission) and concludes on the anchor day. The upper boundary starts 1 day post-anchor point and concludes 14 days afterward.

| (Days)\(Days)<br>Before \ After | 1 | 2 | 3 | 4 | 5 | 6 | 7 | 8 | 9 | 10 | 11 | 12 | 13 | 14 |
| --- | --- | --- | --- | --- | --- | --- | --- | --- | --- | --- | --- | --- | --- | --- |
| 0 | 0.72 | 0.76 | 0.76 | 0.80 | 0.79 | 0.86 | 0.83 | 0.84 | 0.84 | 0.85 | 0.84 | 0.84 | 0.84 | 0.85 |
| 1 | 0.72 | 0.77 | 0.77 | 0.80 | 0.83 | 0.84 | 0.83 | 0.82 | 0.83 | 0.83 | 0.85 | 0.85 | 0.85 | 0.85 |
| 2 | 0.73 | 0.77 | 0.78 | 0.79 | 0.83 | 0.82 | 0.82 | 0.81 | 0.83 | 0.84 | 0.85 | 0.86 | 0.85 | 0.86 |
| 3 | 0.72 | 0.77 | 0.78 | 0.80 | 0.83 | 0.83 | 0.83 | 0.82 | 0.83 | 0.84 | 0.85 | 0.85 | **0.87** | **0.87** |

**Figure 5:**
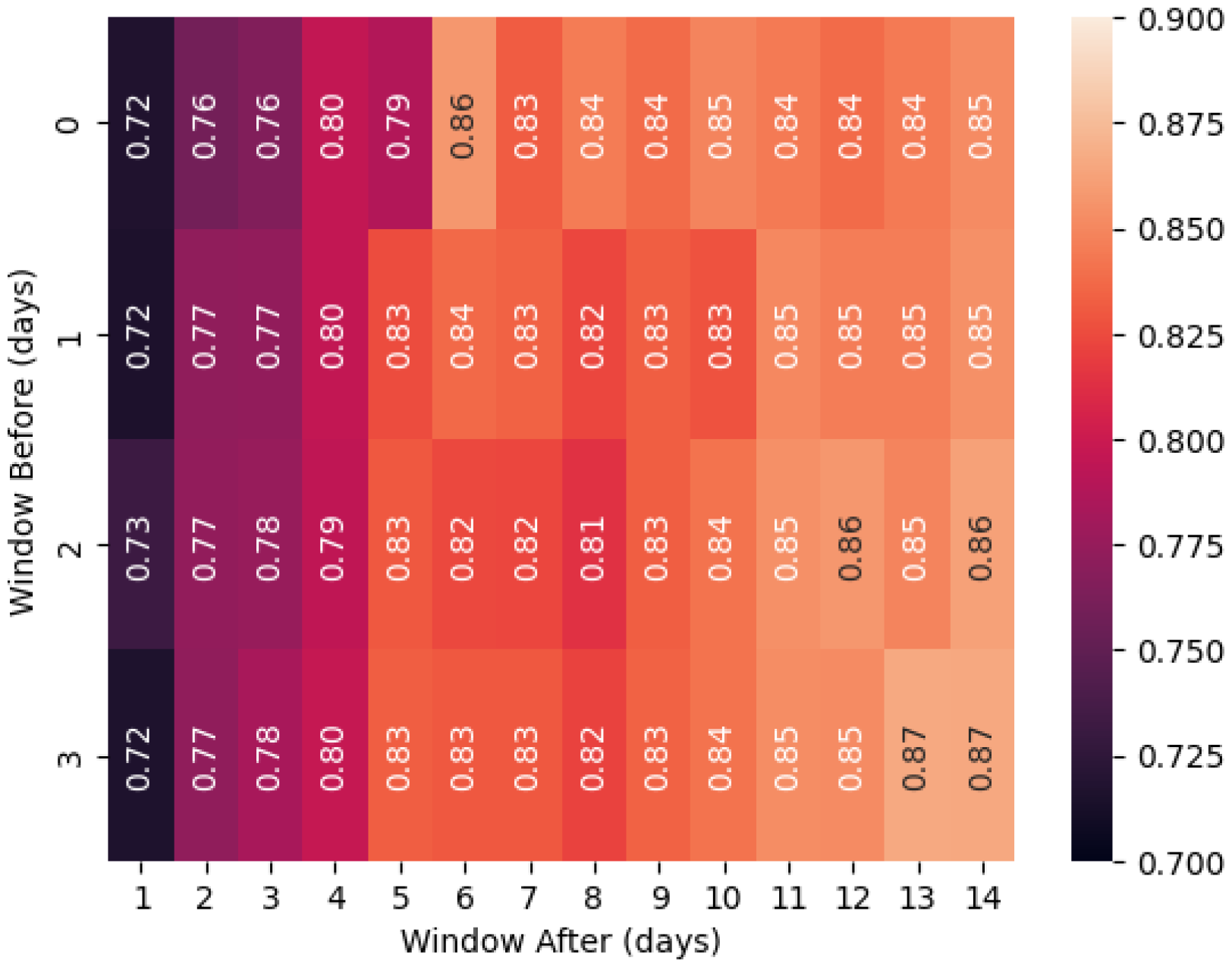
Heatmap displaying AUROC performance across varied data collection windows: Lower boundary initiated 3 days prior to the anchor day (ICU admission) and concluding on the anchor day; upper boundary commencing 1 day post-anchor point and concluding 14 days thereafter. The colour intensity for each cell corresponds to labelled average AUROC values. A legend is provided on the right for reference.

The figure (Figure 5) reveals the impressive performance of EHR-ML even with limited data availability. Notably, the model achieves a good AUROC of 0.72 on day 1, using data from less than 24 hours post-admission. This is comparable to the performance of SAPS II (0.73) built using data collected for more than the first 24 hours. This demonstrates the early predictive power of EHR-ML, potentially enabling timely interventions and improved patient outcomes. Furthermore, as the data window increases, providing more context, EHR-ML’s performance consistently improves, surpassing the benchmark of SAPS II by a significant margin on day 2 at which point there is data of a full 24-hour period. This trend continues as the data window expands, with performance steadily improving due to the model’s ability to capture and utilise temporal information from the time series. This analysis demonstrates EHR-ML’s dual strengths: achieving competitive performance at the very beginning of a patient’s admission and exhibiting continual improvement with longer data availability.

### 3.5. EHR-ML achieves high performance with limited data

To evaluate the impact of sample size on EHR-ML’s performance, we conducted a sample size analysis. We randomly sampled the data from the eICU ICD Cohort, starting with 200 samples and incrementally increasing the size by 100 until reaching 1000. Beyond this point, the sample size increased in steps of 1000, culminating in the full dataset size of 11,146. For each sample size, we performed 5-fold cross-validation and calculated the average and standard error of the AUROC for predicting in-hospital mortality (S4 and figure 6).

**Figure 6:**
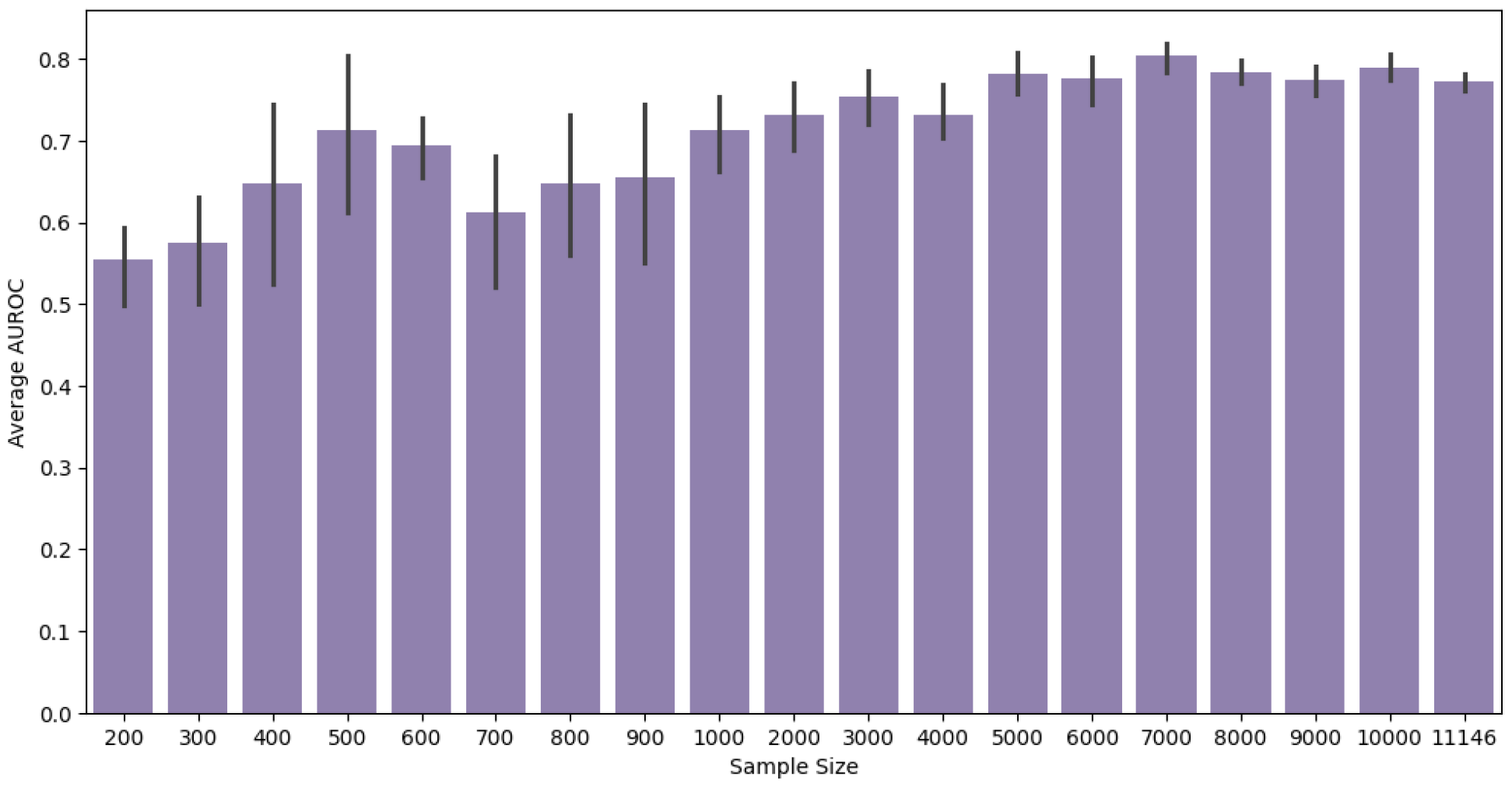
The figure displays a bar plot presenting the average AUROC obtained through 5-fold cross-validation across various sample sizes. Error bars depict standard errors for each sample size. The sample sizes represent the number of episodes used in the analysis, starting from 200 episodes with increments of 100 until 1000. Beyond this, the sample size increases in steps of 1000 until reaching 10000, with the maximum available samples (11146) included at the end.

Figure 6 reveals a substantial initial performance gain as the sample size increases, reaching a plateau of around 500 samples. This suggests that EHR-ML achieves significant performance even with a relatively small dataset, which is remarkable considering the high dimensionality of the data. This can be attributed to the model’s architecture, which divides features into various subsets. This approach allows EHR-ML to effectively handle high-dimensional data without affecting its predictive capabilities. Furthermore, the figure suggests that performance stabilises after 3,000 samples, with minimal variation observed beyond this point. This indicates that 3,000 samples may represent an optimal sample size for achieving stable and reliable results for this analysis using EHR-ML. This insight can be used to improve research design and resource allocation in using this model for practical applications.

### 3.6. Cautionary note on evaluating performance in the face of imbalance

The next EHR-ML analysis explored the effect of class imbalance on standard performance metrics. Class ratio indicates the proportion of various classes in the dataset, like the ratio of positive to negative observations in binary classification. In EHR-based prediction tasks, encountering highly imbalanced classes is common. This analysis aimed to identify metrics better suited for highly imbalanced data and potentially misleading ones. To achieve this, we created datasets with varying class ratios, ranging from a perfect 50-50 balance to a highly skewed 95-5 ratio. We then calculated the average value of various performance measures for each class ratio and tabulated the results in supplementary table S5. Subsequently, a line plot (supplementary figure S3) presenting the trend lines for these measurements is plotted, providing a visual representation of the behaviour of different metrics under different levels of class imbalance.

The supplementary table S5 and supplementary figure S3 provide valuable insights into the impact of class imbalance on various performance metrics. Increasing class imbalance reveals notable discrepancies in metric behavior. AUROC remains relatively stable, while accuracy can be misleadingly inflated, potentially obscuring issues in minority class identification. Conversely, F1 score and MCC exhibit sensitivity to class imbalance, offering a more nuanced evaluation of model performance across all classes. Additionally, MCCF1 [72], combining F1 and MCC, emerges as a promising metric in highly imbalanced data scenarios. These findings underscore the importance of carefully selecting performance metrics for models trained on imbalanced data. Metrics like accuracy may appear intuitive but can be deceptive in such contexts. Instead, prioritizing metrics like MCCF1 provides a more dependable assessment of model performance across diverse classes.

### 3.7. Robust LOS prediction with EHR-ML

Another outcome that is considered in this study is predicting the LOS. Specifically, two separate EHR-ML classifiers were trained to predict whether an ICU admission would exceed 7 and 14 days. To that end, two new target attributes were derived from the existing patient admission records, each corresponding to the respective LOS cut-off. Next, the optimal configuration for both the targets we obtained by running benchmarking analysis. Figure 7 presents the results from this analysis.

**Figure 7:**
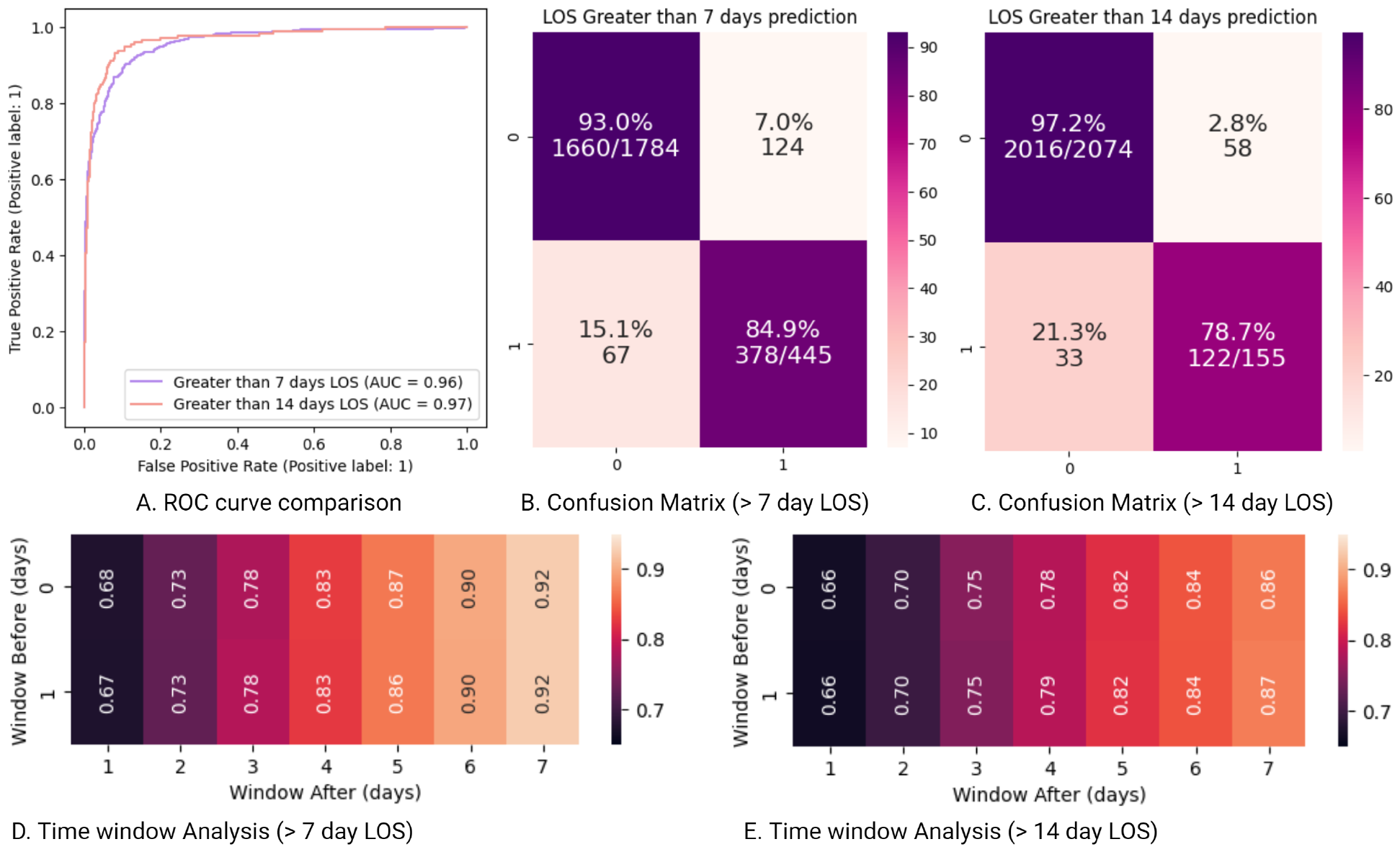
This figure compares the performance of two models in predicting whether a patient’s LOS in the ICU will exceed set durations of 7 and 14 days. A) The plots in Panel A display ROC curves for both the models. This visualisation helps assess the trade-off between sensitivity and specificity across different thresholds. The AUC values are listed for each model in the figure legend along with the corresponding label. B - C) The next two plots in panel B and C show the confusion matrix of the two models. Analysing these values allows us to evaluate the models’ ability to identify both positive and negative cases, as well as the rate of misclassification for each category. In the confusion matrix, the vertical axis represents actual patient LOS status while the horizontal axis corresponds to the predicted outcomes. Both axes use “0” for the negative class (stays below the set duration), and “1” for the positive class (stays over the set duration). Each cell reveals the number and percentage of patients falling into each category, with the total class counts displayed on the diagonal cells. The variation of the colour intensity visually highlights the percentage of observations in each cell. D - E) Panels D and E present heat-maps illustrating the variation in AUROC performance for different data collection windows. Each cell corresponds to an average AUROC value, with colour intensity reflecting the level of performance. The lower boundary of each window starts 1 day before ICU admission (anchor day) and ends on the anchor day, while the upper boundary begins 1 day after the anchor point and concludes 7 days later. A legend for colour interpretation is provided on the right.

The results presented in supplementary tables S6, S7, and the figure 7 illustrate performance of the EHR-ML in predicting LOS exceeding 7 and 14 days. The supplementary table S6 reveals impressive performance for both models in predicting LOS. Notably, the high AUROC values consistently exceeding 0.95 and MCCF1 values over 0.8 indicate excellent discrimination between ICU admissions with long and short stays. Figure 7-A further reinforces these findings through the ROC curves for both models exhibiting excellent coverage. Figures 7-B and 7-C offer visual confirmation of the model’s ability to correctly identify both classes with high confidence. The confusion matrices reveal high percentages for both true positives and true negatives. Furthermore, by examining the diverse time windows and their influence on model performance in supplementary table S7, figures 7-D, and 7-E, researchers can make informed decisions regarding the most suitable window for their specific predictions.

### 3.8. EHR-ML offers a user-friendly interface for clinical outcome prediction from EHR

EHR-ML simplifies model building, performing prediction, running cross-validation evaluation, and performing analysis such as data window analysis, sample size analysis, class ratio analysis, and standardisation analysis. It offers two distinct access points - command line library and a web-portal to promote open research practice. The command line library empowers users with technical expertise to leverage EHR-ML’s functionalities through straightforward installation and execution. This option grants direct control over the analysis process using the command line, enabling customization and integration with existing workflows. In addition, to cater to the users with limited programming experience, the web-portal (refer to figure 8) is made available. It provides an web-based interface for accessing the full spectrum of EHR-ML’s capabilities. Both access points require data to be formatted in a specific manner. EHR-QC [69], our previously developed toolkit, conveniently aids in preprocessing as well as preparing data compatible with EHR-ML. Besides, the simple data representation format required by the utility is easy to construct without relying on any specific tools.

**Figure 8:**
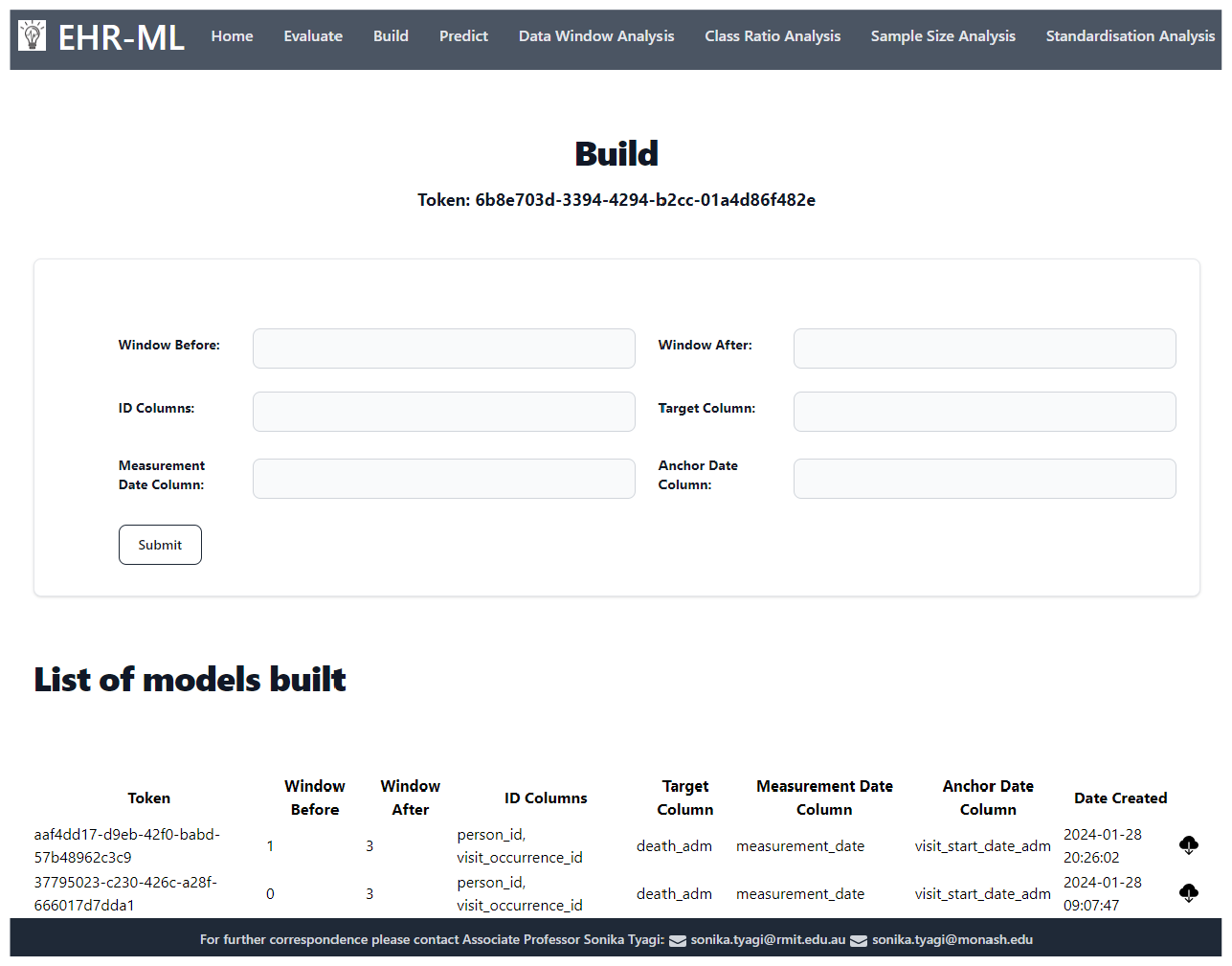
This figure provides a screenshot of the EHR-ML web utility interface. This user-friendly platform enables model building, predicting prediction, running cross-validation, and performing several analyses such as data window analysis, sample size analysis, class ratio analysis, and standardisation analysis. The intuitive interface allows users to upload formatted data and readily conduct various analyses.

## 4. Discussion

The use of machine learning for predicting clinical outcomes shows significant potential in improving healthcare decision-making. However, this field faces limitations that hinder its impact. Our survey reveals a prevalent reliance on expert intuition in constructing predictive models, potentially resulting in suboptimal solutions and hinder automation. Additionally, majority of studies rely on one-time, inflexible codebases, limiting their applicability to various clinical outcomes. The lack of a standardized process impedes the comparison of different studies, hindering the generation of reliable knowledge. Moreover, the use of off-the-shelf models overlooks the complexities of health domain time series measurements and the challenge of class imbalance.

The EHR-ML pipeline emerges as a compelling solution to the challenges encountered in clinical outcome prediction. Moreover, its adaptable interface allows for the customization of data windows, anchor points, and target variables, offering precise control over the modeling process. Additionally, it seamlessly integrates with our EHR-QC preprocessing tool [69], facilitating a streamlined and automated workflow, from data sourcing to outcome modelling. Its innovative approach, characterized by a data-driven methodology and a flexible open-source codebase, holds promise in addressing these hurdles. Through comprehensive benchmarking analysis, we anticipate that the EHR-ML pipeline will streamline the process of knowledge generation and facilitate process automation.One notable strength of the EHR-ML pipeline is its two-layered ensemble modelling approach which offers a proven machine learning model specifically designed for EHR data. This tailored approach enhances the accuracy and reliability of predictive models in healthcare settings. Additionally, the feature engineering functionalities provided by the EHR-ML excels at extracting and leveraging temporal signals from time-series data, a key challenge in this domain. Furthermore, the performance metrics offered by the EHR-ML pipeline provide comprehensive insights into model performance in real-world scenarios, even when dealing with skewed data distributions. This capability ensures that the models can be effectively evaluated and optimized for practical application in healthcare settings.

## 5. Conclusion

EHR-ML represents a notable stride in democratizing and streamlining clinical outcome prediction with EHR data. By eliminating common obstacles and offering a straightforward route to dependable and replicable analyses, it facilitates accessibility. Available both as a command-line interface and a user-friendly web utility, it empowers researchers across various technical proficiencies to conduct predictive modeling for a wide range of clinical outcomes with ease. The open-source nature of the source code encourages community involvement, not only for utilization but also for active contribution. This contribution aims to propel the field forward, fostering reproducibility, comparability, and data-driven optimization in EHR-based clinical outcome studies.

## Data Availability

All data produced in the present study are available upon reasonable request to the authors

## 6. Acknowledgements

AP, NM, GW, and ST acknowledge funding support of Medical Research Future Fund (MRFF) for the SuperbugAI flagship project. YR received Monash Graduate Scholarship for his PhD.

We thank to Jerico Revote from Monash eResearch Centre and William Librata of Alfred Health IT team for their invaluable support to help set up cloud computing infrastructure used in this work.

The authors also extend their sincere appreciation to the open research community responsible for making the following resources available which were instrumental in facilitating the execution of this research; EHR-QC [69], MIMIC IV [73], MIMIC-Extract [70], MIMIC IV to OMOP CDM Conversion (https://github.com/OHDSI/MIMIC), eICU Collaborative Research Database [65], Athena (https://athena.ohdsi.org/), SNOMED (https://www.snomed.org/). Finally, we acknowledge the Python libraries that are an integral part of EHR-ML toolkit. These include scikit-learn, pandas, numpy, psycopg, scipy, matplotlib, xgboost, and lightgbm.

## 7. Code availability

The EHR-ML source code is now accessible to researchers for investigative purposes through the following Git repository: https://gitlab.com/superbugai/ehr-ml. Comprehensive documentation for the utility can also be found at https://ehr-ml-tutorials.readthedocs.io.

### CRediT authorship contribution statement

**Yashpal Ramakrishnaiah:** Formal analysis, Investigation, Visualisation, Writing – review and editing, Software development. **Nenad Macesic:** Investigation, Funding acquisition, Supervision, Writing – review and editing. **Geoffrey I. Webb:** Investigation, Resources, Supervision, Writing – review and editing. **Anton Y. Peleg:** Investigation, Funding acquisition, Resources, Supervision, Writing – review. **Sonika Tyagi:** Conceptualisation, Formal analysis, Investigation, Funding acquisition, Resources, Supervision, Writing – review and editing.

## References

[1] Pranav Rajpurkar, Emma Chen, Oishi Banerjee, and Eric J Topol. AI in health and medicine. Nat. Med., 28(1):31–38, January 2022.

[2] Brian Wahl, Aline Cossy-Gantner, Stefan Germann, and Nina R Schwalbe. Artificial intelligence (AI) and global health: how can AI contribute to health in resource-poor settings? BMJ Global Health, 3(4):e000798, August 2018.

[3] Nina Schwalbe and Brian Wahl. Artificial intelligence and the future of global health. Lancet, 395(10236):1579–1586, May 2020.

[4] Ezekiel J Emanuel and Robert M Wachter. Artificial intelligence in health care: Will the value match the hype? JAMA, 321(23):2281–2282, June 2019.

[5] Min Chen, Xuan Tan, and Rema Padman. Social determinants of health in electronic health records and their impact on analysis and risk prediction: A systematic review. J. Am. Med. Inform. Assoc., 27(11):1764–1773, November 2020.

[6] Nenad Tomašev, Natalie Harris, Sebastien Baur, Anne Mottram, Xavier Glorot, Jack W Rae, Michal Zielinski, Harry Askham, Andre Saraiva, Valerio Magliulo, Clemens Meyer, Suman Ravuri, Ivan Protsyuk, Alistair Connell, Cían O Hughes, Alan Karthikesalingam, Julien Cornebise, Hugh Montgomery, Geraint Rees, Chris Laing, Clifton R Baker, Thomas F Osborne, Ruth Reeves, Demis Hassabis, Dominic King, Mustafa Suleyman, Trevor Back, Christopher Nielson, Martin G Seneviratne, Joseph R Ledsam, and Shakir Mohamed. Use of deep learning to develop continuous-risk models for adverse event prediction from electronic health records. Nat. Protoc., 16(6):2765–2787, May 2021.

[7] Patrick Schwab, Arash Mehrjou, Sonali Parbhoo, Leo Anthony Celi, Jürgen Hetzel, Markus Hofer, Bernhard Schölkopf, and Stefan Bauer. Real-time prediction of COVID-19 related mortality using electronic health records. Nat. Commun., 12(1):1–16, February 2021.

[8] Sujata Khedkar, Priyanka Gandhi, Gayatri Shinde, and Vignesh Subramanian. Deep learning and explainable AI in healthcare using EHR. Deep Learning Techniques for Biomedical and Health Informatics, pages 129–148, 2020.

[9] Awais Ashfaq, Anita Sant’Anna, Markus Lingman, and Sławomir Nowaczyk. Readmission prediction using deep learning on electronic health records. Journal of Biomedical Informatics, 97:103256, 2019.

[10] Matthew B A McDermott, Shirly Wang, Nikki Marinsek, Rajesh Ranganath, Luca Foschini, and Marzyeh Ghassemi. Reproducibility in machine learning for health research: Still a ways to go. Sci. Transl. Med., March 2021.

[11] Adam M Chekroud, Matt Hawrilenko, Hieronimus Loho, Julia Bondar, Ralitza Gueorguieva, Alkomiet Hasan, Joseph Kambeitz, Philip R Corlett, Nikolaos Koutsouleris, Harlan M Krumholz, John H Krystal, and Martin Paulus. Illusory generalizability of clinical prediction models. Science, 383(6679):164–167, January 2024.

[12] Jenny Yang, Andrew A S Soltan, and David A Clifton. Machine learning generalizability across healthcare settings: insights from multi-site COVID-19 screening. npj Digital Medicine, 5(1):1–8, June 2022.

[13] Yuval Barak-Corren, Pradip Chaudhari, Jessica Perniciaro, Mark Waltzman, Andrew M Fine, and Ben Y Reis. Prediction across healthcare settings: a case study in predicting emergency department disposition. npj Digital Medicine, 4(1):1–7, December 2021.

[14] Benjamin A Goldstein, Ann Marie Navar, Michael J Pencina, and John P A Ioannidis. Opportunities and challenges in developing risk prediction models with electronic health records data: a systematic review. J. Am. Med. Inform. Assoc., 24(1):198–208, January 2017.

[15] J E M Vernooij, N J Koning, J W Geurts, S Holewijn, B Preckel, C J Kalkman, and L M Vernooij. Performance and usability of pre-operative prediction models for 30-day peri-operative mortality risk: a systematic review. Anaesthesia, 78(5):607–619, May 2023.

[16] Gary S Collins, Joris A de Groot, Susan Dutton, Omar Omar, Milensu Shanyinde, Abdelouahid Tajar, Merryn Voysey, Rose Wharton, Ly-Mee Yu, Karel G Moons, and Douglas G Altman. External validation of multivariable prediction models: a systematic review of methodological conduct and reporting. BMC Med. Res. Methodol., 14(1):1–11, March 2014.

[17] Sheng-Chieh Lu, Cai Xu, Chandler H Nguyen, Yimin Geng, André Pfob, and Chris Sidey-Gibbons. Machine Learning–Based Short-Term mortality prediction models for patients with cancer using electronic health record data: Systematic review and critical appraisal. JMIR Medical Informatics, 10(3):e33182, March 2022.

[18] Cynthia Yang, Jan A Kors, Solomon Ioannou, Luis H John, Aniek F Markus, Alexandros Rekkas, Maria A J de Ridder, Tom M Seinen, Ross D Williams, and Peter R Rijnbeek. Trends in the conduct and reporting of clinical prediction model development and validation: a systematic review. J. Am. Med. Inform. Assoc., 29(5):983–989, January 2022.

[19] Feng Xie, Han Yuan, Yilin Ning, Marcus Eng Hock Ong, Mengling Feng, Wynne Hsu, Bibhas Chakraborty, and Nan Liu. Deep learning for temporal data representation in electronic health records: A systematic review of challenges and methodologies. Journal of Biomedical Informatics, 126:103980, 2022.

[20] Ben Van Calster, Laure Wynants, Dirk Timmerman, Ewout W Steyerberg, and Gary S Collins. Predictive analytics in health care: how can we know it works? J. Am. Med. Inform. Assoc., 26(12):1651–1654, August 2019.

[21] Effy Vayena, Alessandro Blasimme, and I Glenn Cohen. Machine learning in medicine: Addressing ethical challenges. PLoS Med., 15(11):e1002689, November 2018.

[22] Ran Zhao, Wen Zhang, Zedan Zhang, Chang He, Rong Xu, Xudong Tang, and Bin Wang. Evaluation of reporting quality of cohort studies using real-world data based on RECORD. January 2023.

[23] Gary S Collins and Karel G M Moons. Reporting of artificial intelligence prediction models. Lancet, 393(10181):1577–1579, April 2019.

[24] Viknesh Sounderajah, Hutan Ashrafian, Ravi Aggarwal, Jeffrey De Fauw, Alastair K Denniston, Felix Greaves, Alan Karthikesalingam, Dominic King, Xiaoxuan Liu, Sheraz R Markar, Matthew D F McInnes, Trishan Panch, Jonathan Pearson-Stuttard, Daniel S W Ting, Robert M Golub, David Moher, Patrick M Bossuyt, and Ara Darzi. Developing specific reporting guidelines for diagnostic accuracy studies assessing AI interventions: The STARD-AI steering group. Nat. Med., 26(6):807–808, June 2020.

[25] Po-Hsuan Cameron Chen, Yun Liu, and Lily Peng. How to develop machine learning models for healthcare. Nat. Mater., 18(5):410–414, April 2019.

[26] Norah L Crossnohere, Mohamed Elsaid, Jonathan Paskett, Seuli Bose-Brill, and John F P Bridges. Guidelines for artificial intelligence in medicine: Literature review and content analysis of frameworks. J. Med. Internet Res., 24(8):e36823, August 2022.

[27] Jenna Wiens, Suchi Saria, Mark Sendak, Marzyeh Ghassemi, Vincent X Liu, Finale Doshi-Velez, Kenneth Jung, Katherine Heller, David Kale, Mohammed Saeed, Pilar N Ossorio, Sonoo Thadaney-Israni, and Anna Goldenberg. Do no harm: a roadmap for responsible machine learning for health care. Nat. Med., 25(9):1337–1340, August 2019.

[28] Marzyeh Ghassemi, Tristan Naumann, Peter Schulam, Andrew L Beam, Irene Y Chen, and Rajesh Ranganath. Practical guidance on artificial intelligence for health-care data. The Lancet Digital Health, 1(4):e157–e159, August 2019.

[29] Andre Esteva, Alexandre Robicquet, Bharath Ramsundar, Volodymyr Kuleshov, Mark DePristo, Katherine Chou, Claire Cui, Greg Corrado, Sebastian Thrun, and Jeff Dean. A guide to deep learning in healthcare. Nat. Med., 25(1):24–29, January 2019.

[30] Yikuan Li, Mohammad Mamouei, Gholamreza Salimi-Khorshidi, Shishir Rao, Abdelaali Hassaine, Dexter Canoy, Thomas Lukasiewicz, and Kazem Rahimi. Hi-BEHRT: Hierarchical Transformer-Based model for accurate prediction of clinical events using multimodal longitudinal electronic health records. IEEE J Biomed Health Inform, 27(2):1106–1117, February 2023.

[31] Sarah Alnegheimish, Najat Alrashed, Faisal Aleissa, Shahad Althobaiti, Dongyu Liu, Mansour Alsaleh, and Kalyan Veeramachaneni. Cardea: An open automated machine learning framework for electronic health records. In 2020 IEEE 7th International Conference on Data Science and Advanced Analytics (DSAA). IEEE, October 2020.

[32] Jenna M Reps, Martijn J Schuemie, Marc A Suchard, Patrick B Ryan, and Peter R Rijnbeek. Design and implementation of a standardized framework to generate and evaluate patient-level prediction models using observational healthcare data. J. Am. Med. Inform. Assoc., 25(8):969–975, April 2018.

[33] Chang Hu, Lu Li, Weipeng Huang, Tong Wu, Qiancheng Xu, Juan Liu, and Bo Hu. Interpretable machine learning for early prediction of prognosis in sepsis: A discovery and validation study. Infectious Diseases and Therapy, 11(3):1117–1132, April 2022.

[34] Bernhard Wernly, Behrooz Mamandipoor, Philipp Baldia, Christian Jung, and Venet Osmani. Machine learning predicts mortality in septic patients using only routinely available abg variables: a multi-centre evaluation. International Journal of Medical Informatics, 145:104312, 2021.

[35] Thanakron Na Pattalung and Sitthichok Chaichulee. Comparison of machine learning algorithms for mortality prediction in intensive care patients on multi-center critical care databases. IOP Conf. Ser. Mater. Sci. Eng., 1163(1):012027, August 2021.

[36] Hrayr Harutyunyan, Hrant Khachatrian, David C Kale, Greg Ver Steeg, and Aram Galstyan. Multitask learning and benchmarking with clinical time series data. Scientific Data, 6(1):1–18, June 2019.

[37] Changhee Lee, Jinsung Yoon, and Mihaela van der Schaar. Dynamic-deephit: A deep learning approach for dynamic survival analysis with competing risks based on longitudinal data. IEEE Transactions on Biomedical Engineering, 67(1):122–133, 2020.

[38] Chirag Nagpal, Xinyu Li, and Artur Dubrawski. Deep survival machines: Fully parametric survival regression and representation learning for censored data with competing risks. IEEE Journal of Biomedical and Health Informatics, 25(8):3163–3175, 2021.

[39] Amin Naemi, Thomas Schmidt, Marjan Mansourvar, Ali Ebrahimi, and Uffe Kock Wiil. Quantifying the impact of addressing data challenges in prediction of length of stay. BMC Med. Inform. Decis. Mak., 21(1):1–13, October 2021.

[40] Zhenhui Xu, Congwen Zhao, Charles D Scales, Jr, Ricardo Henao, and Benjamin A Goldstein. Predicting in-hospital length of stay: a two-stage modeling approach to account for highly skewed data. BMC Med. Inform. Decis. Mak., 22(1):110, April 2022.

[41] Richard D Riley, Kym Ie Snell, Joie Ensor, Danielle L Burke, Frank E Harrell, Jr, Karel Gm Moons, and Gary S Collins. Minimum sample size for developing a multivariable prediction model: PART II - binary and time-to-event outcomes. Stat. Med., 38(7):1276–1296, March 2019.

[42] Hong Li, Anja De Waegenaere, and Bertrand Melenberg. The choice of sample size for mortality forecasting: A bayesian learning approach. Insurance: Mathematics and Economics, 63:153–168, 2015. Special Issue: Longevity Nine - the Ninth International Longevity Risk and Capital Markets Solutions Conference.

[43] Jiankang Liu, Xian Xiang Chen, Lipeng Fang, Jun Xia Li, Ting Yang, Qingyuan Zhan, Kai Tong, and Zhen Fang. Mortality prediction based on imbalanced high-dimensional icu big data. Computers in Industry, 98:218–225, 2018.

[44] Sakyajit Bhattacharya, Vaibhav Rajan, and Harsh Shrivastava. ICU mortality prediction: A classification algorithm for imbalanced datasets. AAAI, 31(1), February 2017.

[45] Md Manjurul Ahsan, M A Parvez Mahmud, Pritom Kumar Saha, Kishor Datta Gupta, and Zahed Siddique. Effect of data scaling methods on machine learning algorithms and model performance. Technologies, 9(3):52, July 2021.

[46] Nilay D Shah, Ewout W Steyerberg, and David M Kent. Big data and predictive analytics: Recalibrating expectations. JAMA, 320(1):27–28, July 2018.

[47] Zhongheng Zhang, Lin Chen, Ping Xu, Qing Wang, Jianjun Zhang, Kun Chen, Casey M Clements, Leo Anthony Celi, Vitaly Herasevich, and Yucai Hong. Effectiveness of automated alerting system compared to usual care for the management of sepsis. npj Digital Medicine, 5(1):1–10, July 2022.

[48] Xuan Han, Alexandra Spicer, Kyle A Carey, Emily R Gilbert, Neda Laiteerapong, Nirav S Shah, Christopher Winslow, Majid Afshar, Markos G Kashiouris, and Matthew M Churpek. Identifying High-Risk subphenotypes and associated harms from delayed antibiotic orders and delivery*. Crit. Care Med., 49(10):1694, October 2021.

[49] Christopher W Seymour, Jeremy M Kahn, Christian Martin-Gill, Clifton W Callaway, Donald M Yealy, Damon Scales, and Derek C Angus. Delays from first medical contact to antibiotic administration for sepsis. Crit. Care Med., 45(5):759–765, May 2017.

[50] Joshua P Parreco, Antonio E Hidalgo, Alejandro D Badilla, Omar Ilyas, and Rishi Rattan. Predicting central line-associated bloodstream infections and mortality using supervised machine learning. J. Crit. Care, 45:156–162, June 2018.

[51] Rajsavi S Anand, Paul Stey, Sukrit Jain, Dustin R Biron, Harikrishna Bhatt, Kristina Monteiro, Edward Feller, Megan L Ranney, Indra Neil Sarkar, and Elizabeth S Chen. Predicting mortality in diabetic ICU patients using machine learning and severity indices. AMIA Jt Summits Transl Sci Proc, 2017:310–319, May 2018.

[52] Hamid R Darabi, Daniel Tsinis, Kevin Zecchini, Winthrop F Whitcomb, and Alexander Liss. Forecasting mortality risk for patients admitted to intensive care units using machine learning. Procedia Comput. Sci., 140:306–313, January 2018.

[53] Alistair E W Johnson, Tom J Pollard, and Roger G Mark. Reproducibility in critical care: a mortality prediction case study. In Finale Doshi-Velez, Jim Fackler, David Kale, Rajesh Ranganath, Byron Wallace, and Jenna Wiens, editors, Proceedings of the 2nd Machine Learning for Healthcare Conference, volume 68 of Proceedings of Machine Learning Research, pages 361–376, Boston, Massachusetts, 2017. PMLR.

[54] JL Vincent, R Moreno, J Takala, S Willatts, and others. The SOFA (sepsis-related organ failure assessment) score to describe organ dysfunction/failure. https://www.researchgate.net/profile/Rui-Moreno/publication/14361654_The_SOFA_Sepsis-related_Organ_Failure_Assessment_score_to_describe_organ_dysfunctionfailure_On_behalf_of_the_Working_Group_on_Sepsis-Related_Problems_of_the_European_Society_of_Intensive_Care_Medicine/links/0c960536cf4f20aef4000000/The-SOFA-Sepsis-related-Organ-Failure-Assessment-score-to-describe-organ-dysfunction-failure-On-behalf-of-the-Worpdf, 1996. Accessed: 2021-7-29.

[55] Mervyn Singer, Clifford S Deutschman, Christopher Warren Seymour, Manu Shankar-Hari, Djillali Annane, Michael Bauer, Rinaldo Bellomo, Gordon R Bernard, Jean-Daniel Chiche, Craig M Coopersmith, Richard S Hotchkiss, Mitchell M Levy, John C Marshall, Greg S Martin, Steven M Opal, Gordon D Rubenfeld, Tom van der Poll, Jean-Louis Vincent, and Derek C Angus. The third international consensus definitions for sepsis and septic shock (sepsis-3). JAMA, 315(8):801–810, February 2016.

[56] J R Le Gall, S Lemeshow, and F Saulnier. A new simplified acute physiology score (SAPS II) based on a European/North american multicenter study. JAMA, 270(24):2957–2963, 1993.

[57] Jack E Zimmerman, Andrew A Kramer, Douglas S McNair, and Fern M Malila. Acute physiology and chronic health evaluation (APACHE) IV: hospital mortality assessment for today’s critically ill patients. Crit. Care Med., 34(5):1297–1310, May 2006.

[58] Shinya Iwase, Taka-Aki Nakada, Tadanaga Shimada, Takehiko Oami, Takashi Shimazui, Nozomi Takahashi, Jun Yamabe, Yasuo Yamao, and Eiryo Kawakami. Prediction algorithm for ICU mortality and length of stay using machine learning. Sci. Rep., 12(1):1–9, July 2022.

[59] Imjin Ahn, Hansle Gwon, Heejun Kang, Yunha Kim, Hyeram Seo, Heejung Choi, Ha Na Cho, Minkyoung Kim, Tae Joon Jun, and Young-Hak Kim. Machine Learning-Based hospital discharge prediction for patients with cardiovascular diseases: Development and usability study. JMIR Med Inform, 9(11):e32662, November 2021.

[60] Md. Mahbubur Rahman, Dipanjali Kundu, Sayma Alam Suha, Umme Raihan Siddiqi, and Samrat Kumar Dey. Hospital patients’ length of stay prediction: A federated learning approach. Journal of King Saud University - Computer and Information Sciences, 34(10, Part A):7874–7884, 2022.

[61] Emma Rocheteau, Pietro Liò, and Stephanie Hyland. Temporal pointwise convolutional networks for length of stay prediction in the intensive care unit. In Proceedings of the Conference on Health, Inference, and Learning, CHIL ‘21, page 58–68, New York, NY, USA, 2021. Association for Computing Machinery.

[62] Chien-Hua Chen, Jer-Guang Hsieh, Shu-Ling Cheng, Yih-Lon Lin, Po-Hsiang Lin, and Jyh-Horng Jeng. Early short-term prediction of emergency department length of stay using natural language processing for low-acuity outpatients. The American Journal of Emergency Medicine, 38(11):2368–2373, 2020.

[63] Aya Awad, Mohamed Bader-El-Den, and James McNicholas. Patient length of stay and mortality prediction: A survey. Health Serv. Manage. Res., 30(2):105–120, May 2017.

[64] Alistair E W Johnson, Lucas Bulgarelli, Lu Shen, Alvin Gayles, Ayad Shammout, Steven Horng, Tom J Pollard, Sicheng Hao, Benjamin Moody, Brian Gow, Li-Wei H Lehman, Leo A Celi, and Roger G Mark. MIMIC-IV, a freely accessible electronic health record dataset. Sci Data, 10(1):1, January 2023.

[65] Tom J Pollard, Alistair E W Johnson, Jesse D Raffa, Leo A Celi, Roger G Mark, and Omar Badawi. The eICU collaborative research database, a freely available multi-center database for critical care research. Sci Data, 5:180178, September 2018.

[66] George Hripcsak, Jon D Duke, Nigam H Shah, Christian G Reich, Vojtech Huser, Martijn J Schuemie, Marc A Suchard, Rae Woong Park, Ian Chi Kei Wong, Peter R Rijnbeek, et al. Observational health data sciences and informatics (ohdsi): opportunities for observational researchers. Studies in health technology and informatics, 216:574, 2015.

[67] Maryam Garza, Guilherme Del Fiol, Jessica Tenenbaum, Anita Walden, and Meredith Nahm Zozus. Evaluating common data models for use with a longitudinal community registry. Journal of biomedical informatics, 64:333–341, 2016.

[68] Kevin Donnelly. SNOMED-CT: The advanced terminology and coding system for ehealth. Stud. Health Technol. Inform., 121:279–290, 2006.

[69] Yashpal Ramakrishnaiah, Nenad Macesic, Geoffrey I Webb, Anton Y Peleg, and Sonika Tyagi. EHR-QC: A streamlined pipeline for automated electronic health records standardisation and preprocessing to predict clinical outcomes. J. Biomed. Inform., 147:104509, November 2023.

[70] Shirly Wang, Matthew B. A. McDermott, Geeticka Chauhan, Marzyeh Ghassemi, Michael C. Hughes, and Tristan Naumann. Mimic-extract: a data extraction, preprocessing, and representation pipeline for mimic-iii. In Proceedings of the ACM Conference on Health, Inference, and Learning, CHIL ‘20, page 222–235, New York, NY, USA, 2020. Association for Computing Machinery.

[71] Fei Tony Liu, Kai Ming Ting, and Zhi-Hua Zhou. Isolation forest. In 2008 eighth ieee international conference on data mining, pages 413–422. IEEE, 2008.

[72] Chang Cao, Davide Chicco, and Michael M Hoffman. The mcc-f1 curve: a performance evaluation technique for binary classification. arXiv preprint 2006.11278, 2020.

[73] Alistair Johnson, Lucas Bulgarelli, Tom Pollard, Steven Horng, Leo Anthony Celi, and Roger Mark. Mimic-iv, 2023.

