## Supplementary for "EHR-ML: A generalisable pipeline for reproducible clinical outcomes using electronic health records"

### 8. Supplementary

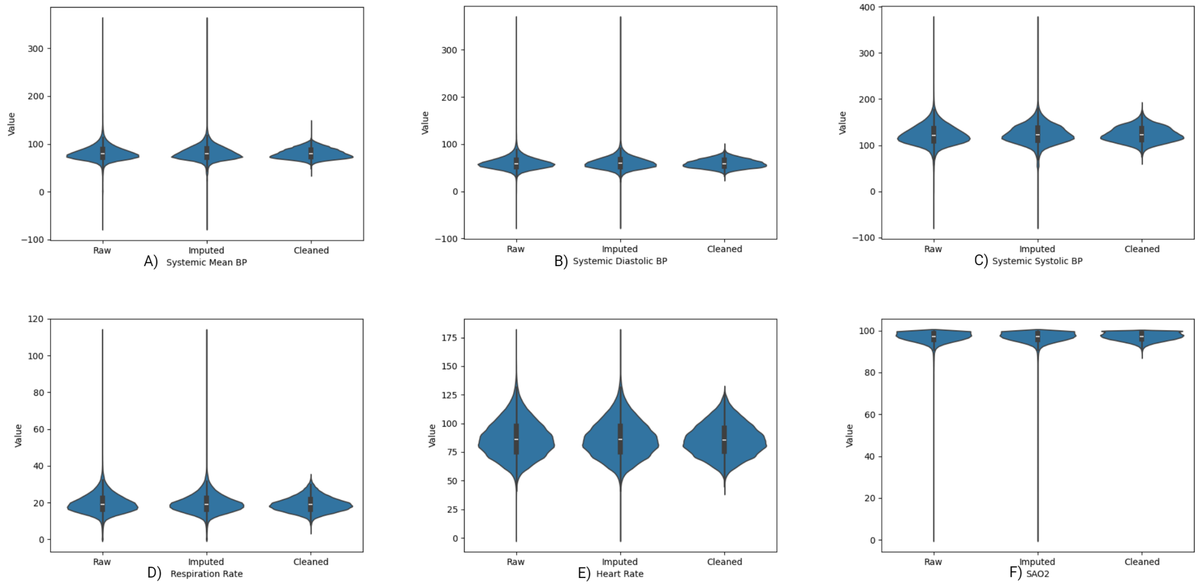


**Figure S1:** This figure summarises the data quality assurance process using EHR-QC. A - F) Each of the six plots ranging from A to F corresponds to a measurement - Systemic Mean BP, Systemic Diastolic BP, Systemic Systolic BP, Respiratory Rate, Heart Rate, and SPO2 respectively. In every plot, the leftmost violins represent the original data, the middle violins show the data after missing values were imputed, and the rightmost violins depict the final cleaned data with outliers removed. The effect of missing data imputation can be observed by comparing the left and middle distributions. Imputation successfully recreates the original distribution, as evidenced by similar variations in the left and middle violin graphs in all the plots (A - F). Similarly, the effect of outlier removal can be observed by comparing the middle and the right distributions. All the six plots (A - F) reveal the impact of outlier removal, where extreme values are eliminated while preserving the core data.

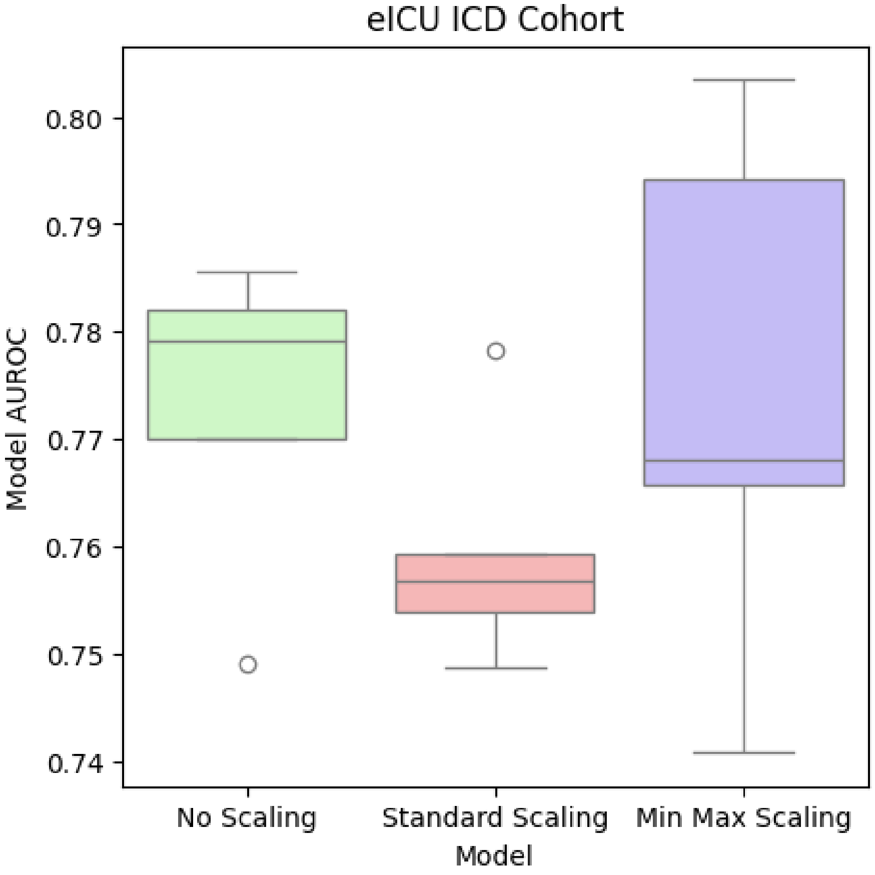


**Figure S2:** The box plots illustrate the model performance, as measured by AUROC, derived from a 5-fold cross-validation. The three plots represent distinct scenarios: the first involves raw (non-scaled) data, the second is for the data scaled using standard scaling resulting in distributions with a mean value of 0 and a standard deviation of 1, and the third plot showcases for the min-max scaled data with a minimum value of 0 and a maximum value of 1. The box plot shows the mean AUROC, upper and lower quartiles, and the minimum and maximum values for all three datasets. This visualisation aims to evaluate whether standardisation is necessary for the specific method or data set under consideration.

| Model | Accuracy | Balanced Accuracy | Average Precision | F1 | AUROC | MCCF1 |
|---|---|---|---|---|---|---|
| XGB (Best Value) | 0.8848 | 0.8918 | 0.5275 | 0.6978 | 0.9614 | 0.7541 |
| LR (Best Value) | 0.9090 | 0.8144 | 0.5203 | 0.6878 | 0.9017 | 0.7442 |
| LGBM (Best Value) | 0.9058 | 0.8973 | 0.5728 | 0.7347 | 0.9662 | 0.7838 |
| MLP (Best Value) | 0.8107 | 0.8585 | 0.4124 | 0.5906 | 0.9478 | 0.6692 |
| ***EHR-ML*** | ***0.9766*** | ***0.9253*** | ***0.8634*** | ***0.9149*** | ***0.9963*** | ***0.9311*** |

**Table S1**

The table presents a comprehensive performance overview, encompassing AUROC, MCCF1, Accuracy, Balanced Accuracy, Average Precision, and F1 for both the EHR-ML ensemble model and its individual constituent models. Notably, EHR-ML demonstrated superior performance compared to the best-performing individual models, across all evaluated dimensions.

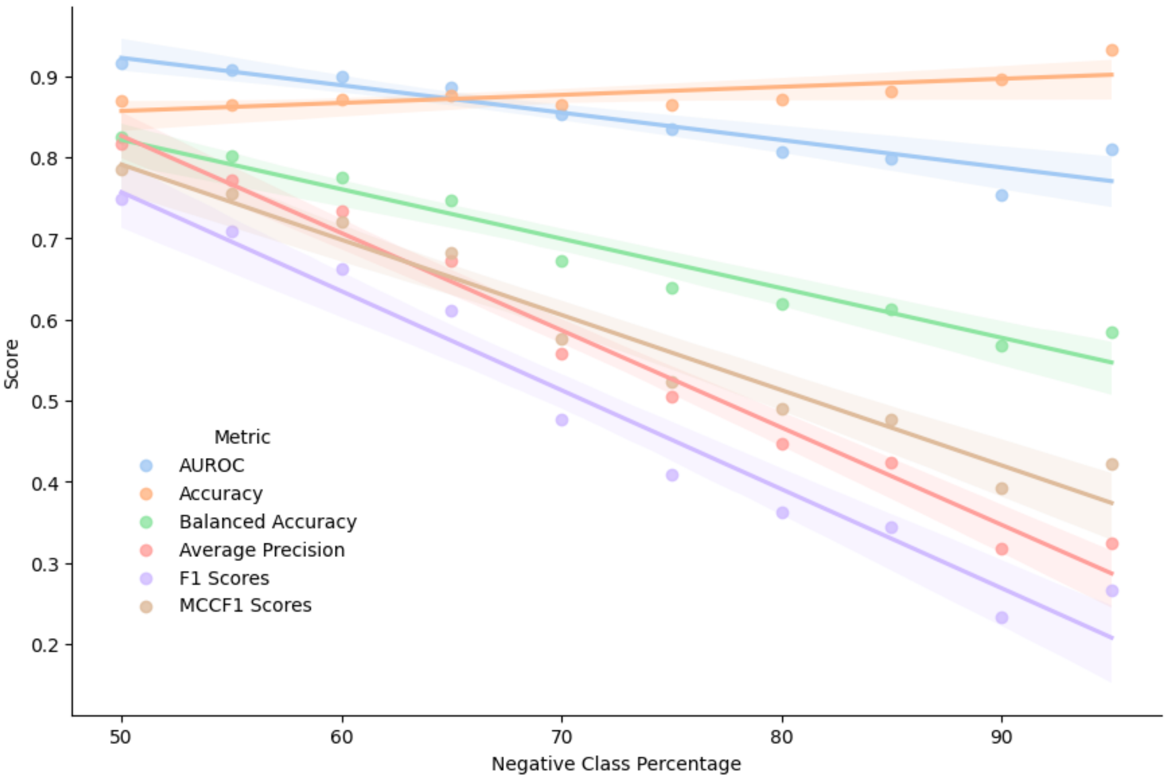


**Figure S3:** Figure depicting the trends of various performance metrics - AUROC, MCCF1, Accuracy, Balanced Accuracy, Average Precision, and F1 - across different class ratios. In this context, "class" pertains to the target variable values predicted by the model, such as "dead" or "alive" when forecasting in-hospital mortality. The dataset is sampled to generate various class ratios, ranging from an equal 50-50 ratio to the most imbalanced dataset with 95% negative observations and 5% positive observations.

| Model | Accuracy | Balanced Accuracy | Average Precision | F1 | AUROC | MCCF1 |
|---|---|---|---|---|---|---|
| LR | 0.9246 | 0.7905 | 0.7595 | 0.7017 | 0.9026 | 0.7596 |
| XGB | **0.9286** | 0.7816 | 0.7848 | 0.7037 | **0.9085** | 0.7636 |
| EHR-ML | 0.9273 | **0.7949** | **0.7970** | **0.7134** | 0.9067 | **0.7696** |

**Table S2**
This table offers a comprehensive comparison of performance metrics for the EHR-ML model and standalone off-the-shelf machine learning models, including XGB and LR. Metrics encompass AUROC, MCCF1, Accuracy, Balanced Accuracy, Average Precision, and F1. In summary, the results display that EHR-ML surpasses off-the-shelf models in its ability to accurately predict clinical outcomes.

| Scaling Method | Mean (AUROC) | Standard Deviation (AUROC) |
|---|---|---|
| ***No Scaling*** | ***0.7732*** | ***0.0146*** |
| Min-Max Scaling | 0.7744 | 0.0249 |
| Standard Scaling | 0.7594 | 0.0112 |

**Table S3**
The table displays the average and standard deviation of the 5-fold cross-validated AUROC for three different data scenarios: raw (non-scaled) data, standard-scaled data with a mean of 0 and a standard deviation of 1, and min-max scaled data with a minimum value of 0 and a maximum of 1. Notably, the results indicate that scaling did not lead to performance gains for EHR-ML for the given dataset.

| Sample Size | Mean (AUROC) | Standard Deviation (AUROC) |
|---|---|---|
| 200 | 0.5547 | 0.0647 |
| 300 | 0.5759 | 0.0907 |
| 400 | 0.6487 | 0.1436 |
| 500 | 0.7129 | 0.1257 |
| 600 | 0.6939 | 0.0497 |
| 700 | 0.6132 | 0.1059 |
| 800 | 0.6483 | 0.1115 |
| 900 | 0.6551 | 0.1162 |
| 1000 | 0.7123 | 0.0595 |
| 2000 | 0.7320 | 0.0566 |
| 3000 | 0.7540 | 0.0421 |
| 4000 | 0.7310 | 0.0429 |
| 5000 | 0.7826 | 0.0322 |
| 6000 | 0.7760 | 0.0380 |
| 7000 | 0.8035 | 0.0229 |
| 8000 | 0.7846 | 0.0184 |
| 9000 | 0.7754 | 0.0231 |
| 10000 | 0.7891 | 0.0190 |
| 11146 | 0.7732 | 0.0147 |

**Table S4**
A table showing the mean AUROC and its standard deviation from a 5-fold cross-validation for different sample sizes obtained from sample-size analysis.

| Ratio | AUROC | Accuracy | Average Precision | Balanced Accuracy | F1 | MCCF1 |
|---|---|---|---|---|---|---|
| 50-50 | **0.9152** | 0.8698 | **0.8161** | **0.8251** | **0.7480** | **0.7853** |
| 55-45 | 0.9071 | 0.8652 | 0.7710 | 0.8016 | 0.7086 | 0.7544 |
| 60-40 | 0.8992 | 0.8709 | 0.7338 | 0.7743 | 0.6630 | 0.7208 |
| 65-35 | 0.8864 | 0.8764 | 0.6728 | 0.7474 | 0.6118 | 0.6819 |
| 70-30 | 0.8529 | 0.8646 | 0.5573 | 0.6717 | 0.4763 | 0.5761 |
| 75-25 | 0.8355 | 0.8652 | 0.5041 | 0.6394 | 0.4081 | 0.5237 |
| 80-20 | 0.8072 | 0.8705 | 0.4468 | 0.6187 | 0.3627 | 0.4896 |
| 85-15 | 0.7978 | 0.8817 | 0.4233 | 0.6121 | 0.3446 | 0.4770 |
| 90-10 | 0.7537 | 0.8962 | 0.3172 | 0.5681 | 0.2333 | 0.3928 |
| 95-05 | 0.8097 | **0.9325** | 0.3239 | 0.5841 | 0.2667 | 0.4222 |

**Table S5**
The table presents various performance metrics, including AUROC, MCCF1, Accuracy, Balanced Accuracy, Average Precision, and F1, across different class ratios. The dataset is sampled to create varying class ratios, spanning from an equal 50-50 ratio to the most imbalanced dataset with 95% negative observations and 5% positive observations.

| Target | AUROC | Accuracy | Average Precision | Balanced Accuracy | F1 | MCCF1 |
|---|---|---|---|---|---|---|
| LOS $>$ 7 days | 0.9593 | 0.9178 | 0.8993 | 0.8732 | 0.8101 | 0.8408 |
| LOS $>$ 14 days | 0.9693 | 0.9636 | 0.8362 | 0.8516 | 0.7546 | 0.8030 |

**Table S6**
This table summarises the performance metrics for predicting LOS exceeding 7 and 14 days. The metrics include AUROC, MCCF1, Accuracy, Balanced Accuracy, Average Precision, and F1. They are obtained using averaging over 5-fold cross-validation performed for predicting LOSgreater than 7 and 14 days. These metrics provide a comprehensive evaluation of the model's ability to accurately classify patients based on their LOS.

| | LOS > 7 days | | | | | | | LOS > 14 days | | | | | | |
|---|---|---|---|---|---|---|---|---|---|---|---|---|---|---|
| (Days)\(Days) Before \ After | 1 | 2 | 3 | 4 | 5 | 6 | 7 | 1 | 2 | 3 | 4 | 5 | 6 | 7 |
| 0 | 0.68 | 0.73 | 0.78 | 0.83 | 0.87 | 0.90 | **0.92** | 0.66 | 0.70 | 0.75 | 0.78 | 0.82 | 0.84 | 0.86 |
| 1 | 0.67 | 0.73 | 0.78 | 0.83 | 0.86 | 0.90 | **0.92** | 0.66 | 0.70 | 0.75 | 0.79 | 0.82 | 0.84 | **0.87** |

**Table S7**

This table summarises average AUROC scores for predicting LOS exceeding 7 and 14 days. Each cell in the table represents a different data collection window, defined by its lower and upper boundaries. The lower boundary starts 1 day before ICU admission (anchor day) and ends on the anchor day, while the upper boundary begins 1 day after admission and ends 7 days later.
